# Treating intrusive memories after trauma in healthcare workers: a Bayesian adaptive randomised trial developing an imagery-competing task intervention

**DOI:** 10.1101/2022.10.06.22280777

**Authors:** Varsha Ramineni, Philip Millroth, Lalitha Iyadurai, Thomas Jaki, Jonathan Kingslake, Julie Highfield, Charlotte Summers, Michael B. Bonsall, Emily A. Holmes

## Abstract

Intensive care unit (ICU) staff continue to face recurrent work-related traumatic events throughout the COVID-19 pandemic. Intrusive memories (IMs) of such traumatic events comprise sensory image-based memories. Harnessing research on preventing IMs with a novel behavioural intervention on the day of trauma, here we take critical next steps in developing this approach as a treatment for ICU staff who are already experiencing IMs days, weeks, or months post-trauma. To address the urgent need to develop novel mental health interventions, we used Bayesian statistical approaches to optimise a brief imagery-competing task intervention to reduce the number of IMs. We evaluated a digitised version of the intervention for remote, scalable delivery. We conducted a two-arm, parallel-group, randomised, adaptive Bayesian optimisation trial. Eligible participants worked clinically in a UK NHS ICU during the pandemic, experienced at least one work-related traumatic event, and at least three IMs in the week prior to recruitment. Participants were randomised to receive immediate or delayed (after four weeks) access to the intervention.

Primary outcome was the number of IMs of trauma during week 4, controlling for baseline week. Analyses were conducted on an intention-to-treat basis as a between-group comparison. Prior to final analysis, sequential Bayesian analyses were conducted (*n*=20,23,29,37,41,45) to inform early stopping of the trial prior to the planned maximum recruitment (*n*=150). Final analysis (*n*=75) showed strong evidence for a positive treatment effect (Bayes factor, BF=1.25 × 10^6^): the immediate arm reported fewer IMs (median=1, IQR=0-3) than the delayed arm (median=10, IQR=6-16.5). With further digital enhancements, the intervention (*n*=28) also showed a positive treatment effect (BF=7.31). Sequential Bayesian analyses provided evidence for reducing IMs of work-related trauma for healthcare workers. This methodology also allowed us to rule out negative effects early, reduced the planned maximum sample size, and allowed evaluation of enhancements. Trial Registration NCT04992390 (www.clinicaltrials.gov).

## INTRODUCTION

Throughout the COVID-19 pandemic, frontline healthcare workers have been repeatedly exposed to potentially psychologically traumatic events, such as untimely and excess deaths of patients. After trauma, it is common to experience intrusive memories (IMs) of the event. These are emotional, sensory, and primarily visual memories (mental imagery) of the traumatic event that intrude repeatedly into mind, comprising a core clinical feature of post-traumatic stress disorder (PTSD) [1]. Anecdotal examples of an IM include a vivid mental image of the eyes of a young patient dying while resuscitation fails; or an image of an ambulance stretcher bearing a former colleague. IMs can be distressing and disruptive. Even before COVID-19, emergency-room nurses reported high levels of IMs of work-related trauma [2], and meta-analysis estimates show that healthcare workers are twice as likely to develop PTSD compared to the general public [3]. Due to the COVID-19 pandemic, around 40% of healthcare workers in UK hospitals reported a level of symptoms consistent with a diagnosis of PTSD as of June/July 2020 [4] - five times higher than in 2015 [5]. Reports of mental health disorders in healthcare workers increased further to 64% in winter 2020 of the pandemic [6].

The urgent need for scalable approaches to support the mental health of frontline healthcare workers, such as Intensive Care Unit (ICU) staff, was highlighted early in the pandemic [7]. To address this, novel approaches are needed for this population. Given their high workload demands, a brief and flexible intervention approach would be beneficial. Moreover, the nature of the trauma exposure facing healthcare workers is not single trauma but repeated and ongoing through the pandemic. Harnessing research on preventing IMs with a novel behavioural intervention on the day of trauma [8], here we take critical next steps to develop this approach as a treatment for ICU staff who are already experiencing IMs days, weeks or months post-trauma. The new intervention approach to reduce IMs here aims to be readily repeatable for different traumas as well as brief, flexible and low stigma.

An imagery-competing task intervention approach (which includes computer gameplay) to prevent IMs was developed from insights from cognitive neuroscience and experimental research [9, 10] by taking a mechanistically informed single-symptom approach [11]. The aim was to offer a new low-intensity intervention post-trauma. The intervention included a memory reminder cue plus playing a computer game with high visuospatial demands (Tetris^®^) using mental rotation, theorised to disrupt the consolidation of sensory elements of the trauma memory. More specifically, the intervention exploits principles of working memory theory (i.e., limited capacity to process similar cognitive information simultaneously), and the malleability of memory to updating, by using a competing cognitive task to interfere with mental imagery-based (mainly visuospatial) trauma memory [12]. The disruption of visuospatial processing while memory is being stored or updated should render the memory less liked to be triggered, i.e. from becoming intrusive [13].

A first proof-of-concept translation study from lab to clinic included patients in the emergency department (ED) within 6 hours of a motor vehicle accident [8]. It was predicted that if the intervention was administered in the first hours after a traumatic event, the number of IMs would be reduced. The randomised controlled trial (RCT) compared the imagery-competing task intervention with an attention-placebo control. Results indicated the efficacy of the intervention in that there were fewer IMs reported in the week post-intervention.

Participants found the single session intervention easy, helpful, and minimally distressing. Similar findings were found in a subsequent study in the ED [14] including more trauma types than motor vehicle accidents. Further, addressing a previous limitation, results showed the reduction in IMs at 1 week persisted to 1 month. A similar intervention approach with mothers after traumatic childbirth showed an effect reducing IMs [15].

While showing promise for the *prevention* of IMs after trauma [8, 12, 14, 15], critically we also need interventions for the treatment of *established* IMs i.e. rather than on the day of trauma, treatment delivery when a longer time has elapsed (days, weeks or months later) for individuals already experiencing IMs. Accordingly, to adapt the imagery-competing task intervention we drew on insights from memory reconsolidation studies on older established memories [10, 12, 16, 17]. In laboratory [18] and case studies [19, 20] we adapted parameters of the intervention (e.g. timings) to promote a reduction of established IMs (Anemone™).

We digitised intervention delivery procedures [21] so that they could be administered remotely due to contagion risk during the COVID-19 pandemic. In collaboration with healthcare workers with lived experiences of IMs, the tailored intervention was piloted in the pandemic [22].

In the current trial, we sought to optimise and evaluate a brief digital imagery-competing task intervention [8, 18] to reduce the number of IMs of work-related traumatic events for ICU staff. The imagery-competing task intervention consisted of a brief reminder cue to a specific intrusive memory, followed by playing the computer game Tetris^®^ for 20 minutes with mental rotation. The first session was guided by a researcher. Thereafter the intervention could be used self-guided, i.e. was repeatable. Participants in the present trial were already experiencing IMs, with many facing ongoing trauma exposure. This is the first trial to evaluate the brief imagery-competing task intervention for traumatic events that could have taken place days, weeks or months ago, for healthcare workers, and using a remotely delivered digitised version of the intervention on i-spero^®^. The primary outcome diary was four weeks after the guided intervention session.

Given the rapid need for novel approaches during a pandemic, it was advantageous to evaluate the intervention, both swiftly and robustly. However, RCTs are notoriously lengthy, spanning several years before an intervention is optimised and evaluated. The COVID-19 pandemic has led to difficulties with participant recruitment and testing, underscoring the need to minimise sample sizes where possible [23]. Our solution to the challenge of being able to assess evidence rapidly while upholding, and arguably even improving, the standards of RCTs was to apply advances in Bayesian statistical methodology. Bayesian methods provide powerful tools for inference (see ‘Statistical Analysis’), and have been applied widely in medical research, e.g. for SARS-CoV-2 virus [24]. However, reporting of frequentist null hypothesis significance testing is still the prevailing norm in clinical trials [25].

The sequential Bayesian design used here has previously been recommended for vaccine development [26] and used in COVID-19 trials to lower required sample sizes without loss of scientific integrity [23]. For example, in early-phase COVID-19 vaccine trials, aspects of treatment such as dosage were modified based on early evidence, allowing later confirmatory trials to test optimised versions of a treatment [23, 27]. Compared to traditional designs, a sequential Bayesian approach typically requires 50% to 70% smaller samples to conclude the presence of an effect and has the same or lower rate of false inference [28]. Using a sequential Bayesian approach, with the ability to quantify and track evidence over time, allowed for continuous learning from the data to guide decision making (such as early stopping and optimising the intervention). It provided the possibility of taking early action if we saw evidence of a negative effect (greater number of IMs), as well as to act on evidence of a positive treatment effect (fewer IMs) (see ‘Results’).

In sum, we aimed to optimise a brief digital imagery-competing task intervention to help reduce the number of IMs of work-related trauma being experienced by ICU staff. To this end, we used Bayesian statistical methodology to optimise trial design and guide decisions. Taken all together, our aim was that this intervention and the Bayesian methodology would help address the need for accelerated mental health treatment development for healthcare staff working during the COVID-19 pandemic.

## MATERIAL AND METHODS

### Study design and participants

We conducted a two-arm, parallel-group, randomised, adaptive Bayesian optimisation trial of a remotely delivered digital intervention. Ethical approval for the trial was granted by the Wales Research Ethics Committee (Wales REC 6, 21/WA/0173). The trial was registered prospectively at Clinical.Trials.gov (CTR: NCT04992390). The study protocol was added to a public depository (osf.io/2xn5m), and the trial had a data monitoring committee (DMC).

The study was advertised via email and Twitter directly from the Intensive Care Society to its membership network, mailing list, and existing social media followers, supplemented by advertisements through Facebook. Advertisements contained a link to the study website: https://www.p1vital-gains.com/, which included a summary of study information, a video explaining IMs, and a participant information sheet.

Eligible participants were adults aged 18 years or older, who worked in a clinical role in an NHS ICU or equivalent during the COVID-19 pandemic (e.g. as a member of ICU staff or deployed to work in the ICU during the pandemic), who had experienced at least one traumatic event related to their work (meeting criterion A of the DSM-5 criteria for PTSD: “exposure to actual or threatened death, serious injury, or sexual violence” by “directly experiencing the traumatic event(s)” or “witnessing, in person, the event(s) as it occurred to others”), had IMs of the traumatic event(s), and had experienced at least three IMs in the week prior to screening. Further, participants had internet access; were willing and able to be contacted by the research team during the study period, and to provide informed consent and able to complete study procedures, read, write, and speak English. Exclusion criteria were having fewer than three IMs during the baseline week after informed consent (i.e. the run-in week on Fig. 1). We did not exclude those undergoing other treatments for PTSD or its symptoms, so the study was as inclusive as possible to meet the challenges that ICU staff were facing during the COVID-19 pandemic. Written informed consent was obtained before participation (using an electronic signature via email).

**Fig. 1.**
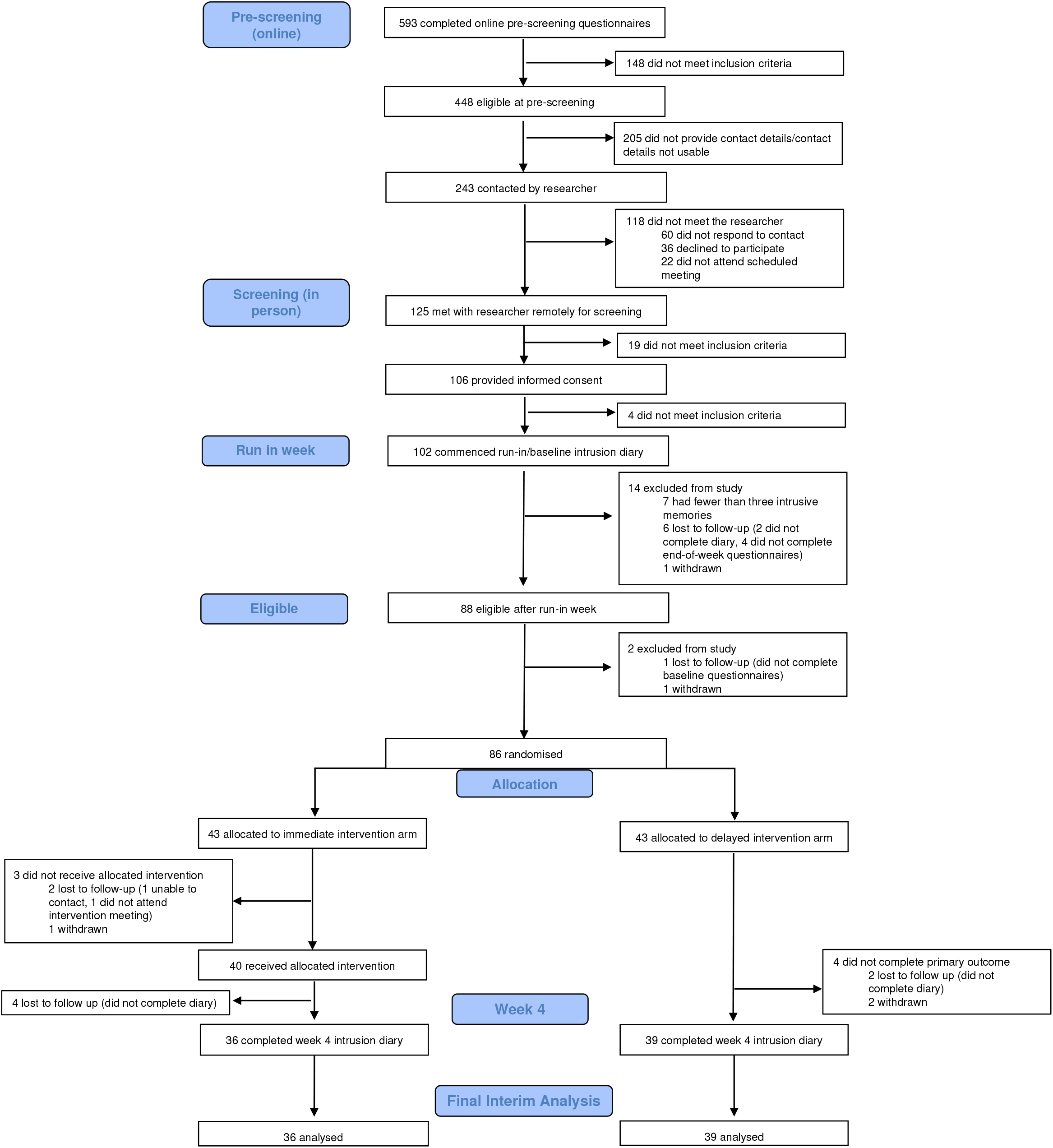
Trial Profile. CONSORT diagram showing enrolment, allocation, and the analysis populations

### Randomisation and masking

Participants were randomly assigned (1:1) using a remote, secure web-based clinical research system (P1vital^®^ ePRO) to either immediate intervention arm (immediate access to the brief digital imagery-competing task intervention plus symptom monitoring for four weeks) or delayed intervention arm (usual care for four weeks followed by access to the intervention plus symptom monitoring for four weeks).

Randomisation occurred following the baseline week, and after baseline questionnaires (see protocol; osf.io/2xn5m). The allocation sequence was computer-generated consecutively using minimisation. Minimisation allocated the first participant to either arm, thereafter allocation was preferential to the arm with the fewest participants to minimise the difference in group sizes. The randomisation allocation percentage was originally set to 66% and altered to 85% after 61 participants were randomised (to ensure balance of groups after early stopping decision, see ‘Results’).

Participants were blinded to group allocation. The statistician who conducted the interim analyses (VR), and researchers who contacted participants and facilitated the conduct of the task intervention were not blinded to group allocation. Outcome assessments were masked to group allocation since they were self-reported by participants in the digital platform.

### The intervention

Straight after randomisation, participants in the immediate intervention arm gained access to the digital imagery-competing task intervention with symptom monitoring of IMs for four weeks. The intervention was delivered on a secure web platform (i-spero®) via smartphone, tablet, or computer. Participants had an initial researcher-guided session (approximately one hour, via Microsoft Teams) and thereafter used i-spero® in a self-directed manner (approximately 25 minutes; with the option for support). The researcher-guided session consisted of step-by-step instructions, animated videos and multiple-choice questions.

Participants were instructed to list their IMs by typing a brief description. They selected one IM from their list, and very briefly brought the image to mind. After instructions on playing the computer game Tetris® using mental rotation, they played for 20 minutes. Finally, they were instructed on monitoring IMs in i-spero® and encouraged to use the intervention to target each memory on their list.

The brief digital intervention on i-spero^®^ and P1vital^®^ ePRO are owned and manufactured by P1vital Products Ltd. Tetris^®^ has been licensed for use within i-spero^®^ from The Tetris Company. P1vital^®^ ePRO, i-spero^®^ and the brief digital intervention have been developed following a formal computerized system validation methodology which complies with Good Clinical Practice, FDA 21CFR Part 11 and ISO13485 Quality Management System.

### Assessments

#### Baseline

After informed consent, participants completed a daily IM diary online for seven days (baseline week, day zero to six i.e. run-in week on Fig.1) to record the number of IMs of traumatic event(s). This diary was adapted from previous studies [8, 18] for digital delivery using P1vital^®^ ePRO. Participants were asked “Have you had any intrusive memories today?” and if answered ‘yes’ selected how many, prompted by email/SMS once daily. Those who reported three or more IMs during baseline week and completed baseline questionnaires (sent after baseline week to those meeting the study entry criteria, see Fig. 1 and Supplementary Table 1) were randomised. The total number of IMs of traumatic event(s) recorded during the baseline week is used as a baseline covariate when modelling.

#### Primary outcome

During week 4, participants in both arms were asked to again complete the daily IM diary for seven days (i.e. from day 22 to 28, where day one is the guided session in immediate arm/equivalent timeframe in delayed arm) to record the number of IMs of traumatic event(s); The primary outcome measure was the total number of IMs recorded by participants in this daily IM diary in week 4.

The outcome measure was derived from diaries used in clinical practice [29], laboratory [18] and patient studies [8]. Positive relationships between diary IMs and the Impact of Events Intrusion subscale indicates convergent validity with PTSD symptoms [30]. Count data provides greater sensitivity than questionnaires with finite categories, with no upper bound. Daily completion can reduce retrospective recall biases of completing measures after one week. Service users report the diary is straightforward with typically good adherence and limited missing data [30]. Remote and digital completion of the diary here meant it was assessor blinded.

The clinical meaning of a change in score may depend on the trauma population, as single-event trauma incurs fewer IMs than repeated trauma. For a PTSD diagnosis, the Clinician-Administered PTSD Scale for DSM–5 (CAPS-5) [31] requires at least two IMs over the past month. The CAPS-5 maximum score is ‘daily’, and reducing this to ‘once-or-twice a week’/’never’ (CAPS-5 mild-minimum) represents a clinically meaningful outcome target [32].

#### Safety

Adverse events were monitored through a standardised question (“have you experienced any untoward medical occurrences or other problems?”) at week 4 and week 8, as well as through any spontaneous reports from participants at any time point during the study.

#### Other outcomes

The present article focuses solely on the sequential Bayesian analyses on the primary outcome measure. A standard analysis (using frequentist statistics) of the final study population including secondary outcome measures will be reported elsewhere [33] (CTR: NCT04992390).

## Statistical analysis

Informed by power estimates based on an effect size of *d*=0.63 (based on pooled information from three previous related RCTs [8, 14, 15]), for the primary outcome, we planned to recruit up to 150 participants, with the potential to end recruitment earlier based on interim analyses. Therefore, we employed a sequential Bayesian design with maximal sample size [28, 34]..

This allowed for interim analyses to guide decision-making, such as when to adjust aspects of the intervention to optimise its effect, and when sufficient evidence has been collected to end the optimisation trial, and proceed to a follow-up pragmatic RCT to test the clinical effectiveness of the optimised intervention.

The fundamental idea to a Bayesian approach is simple [35]: the parameters that we are trying to estimate are treated as random variables with distributions that represent our initial beliefs and uncertainty. After observing data, initial beliefs can be updated with the new information to get improved beliefs. This contrasts with the prevailing ‘frequentist’ statistical frameworks where these parameters are fixed, and probabilities are seen as long-run frequencies generated by some unknown process. The principal outcome of fitting a Bayesian model is the posterior distribution: a probability distribution that indicates how probable particular parameter values are, given the prior distribution (representing initial beliefs) and the observed data. In a Bayesian model, the 95% credibility interval states that there is 95% chance that the true population value falls within this interval.

All analyses were completed in R (version 4.1.2) on an intention-to-treat basis. We fitted a Bayesian model, where the primary outcome (intrusive memory count) was modelled using a using a Poisson linear mixed model. The baseline number of IMs, and treatment assignment were fitted as fixed effects with a random intercept effect for participant. As daily IM diary data used to calculate the primary outcome was collected sequentially over time (baseline or week 4), we used time series methods and an expectation-maximisation algorithm [36] to impute missing values. We present the median and interquartile range (IQR) to account for outliers and skewed primary outcome data (Fig 2. and Supplementary Fig 1); other summary statistics such as the mean and standard deviation (SD) can be found in Supplementary Table 2. The posterior mean of the treatment assignment parameter and the associated 95% credible interval will also be presented. The Supplementary Information provides full details regarding software, model assumptions, priors, missing data, model fit (Supplementary Fig. 2 and 3, Supplementary Table 3), and sensitivity analyses (Supplementary Fig. 4-7).

**Fig. 2.**
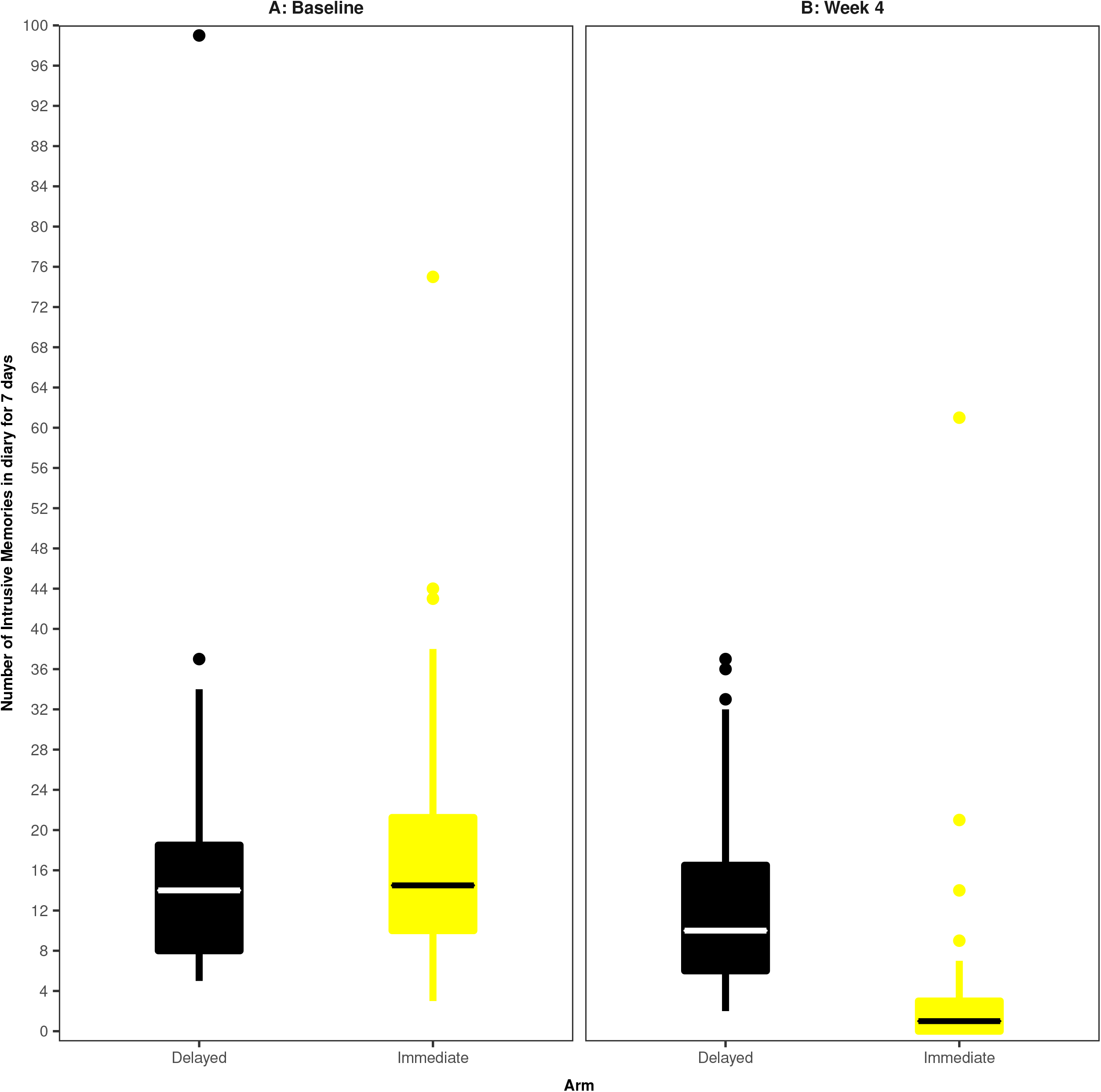
Boxplots for intrusive memory (IM) measures. The midline of the boxplot is the median value, with the upper and lower limits of the box being the third and first quartile (75th and 25th percentile), and the whiskers covering 1.5 times the IQR. The dots depict outliers (each dot represents one subject that departed more than 1.5 times the IQR above the third quartile and below the first quartile). All outliers are included in this figure. **(*A*) Baseline measure for each arm**. Number of IMs of traumatic events recorded by participants in a brief daily online intrusive memory diary for 7 days during the baseline week (i.e. run-in week) for both arms (black = delayed arm (control); *n* = 39: usual care for four weeks; yellow = immediate arm; *n* = 36: immediate access to the intervention following the baseline week: the intervention consisted of a cognitive task involving a trauma reminder-cue plus Tetris computer gameplay using mental rotation plus symptom monitoring), showing that the two arms did not differ at baseline (i.e., before the intervention was provided to the immediate arm. **(*B*) The primary outcome measure for each arm**. Number of IMs of traumatic events recorded by participants in a brief daily online intrusive memory diary for 7 days during week 4 for each arm (black = delayed arm (control); *n* = 39: usual care for four weeks; yellow = immediate arm; *n* = 36: immediate access to the intervention following the baseline week: the intervention consisted of a cognitive task involving a trauma reminder-cue plus Tetris^®^ computer gameplay using mental rotation plus symptom monitoring), showing that the immediate arm had fewer IMs at week 4 compared to the delayed arm and that the number of IMs for the immediate arm decreased between the baseline week and week 4.

**Fig. 3.**
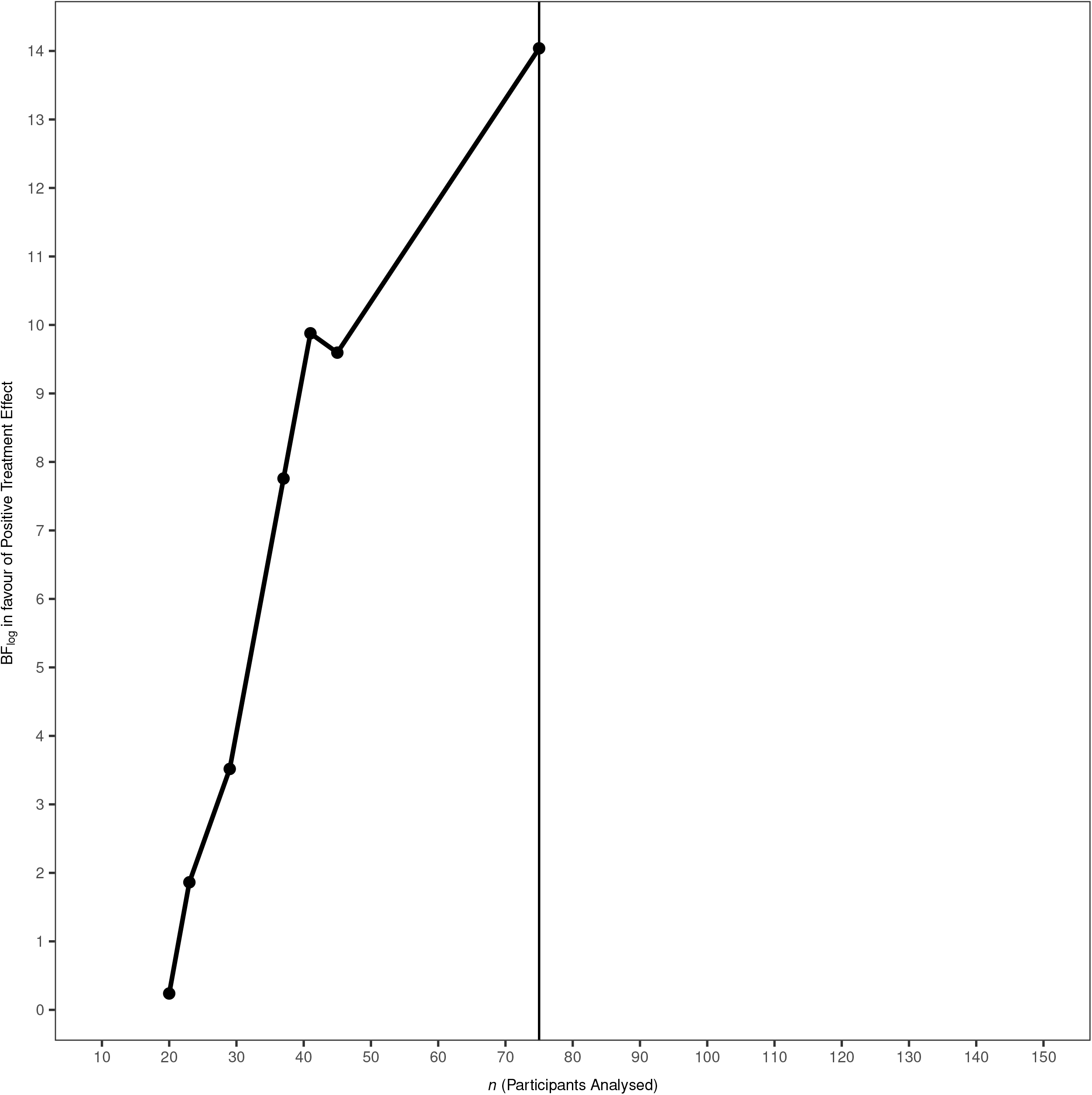
Evidence progression. Sequential Bayesian analyses showed that the evidence (Bf_log_, i.e., the logarithmically scaled Bayes factor) in favour of the hypothesis that the intervention caused a positive treatment effect rapidly increased from the point of the first Bayesian analysis (*n* = 20, raw Bayes factor = 1.27) to the point where on consultation with the DMC the study was concluded (dotted line: *n* = 45, raw Bayes factor = 14,700) pending a round of testing of digital optimisations. The study completed with 86 randomised subjects, 75 of which were analysed (black line), thus saving 64 subjects compared to the original estimated sample size (*n* = 150).

In Bayesian hypothesis testing, a metric known as a Bayes Factor (BF) [35] provides a continuous measure quantifying how well a hypothesis predicts the data relative to a competing hypothesis. Conventional significance tests using p-values do not provide any information about the alternative hypothesis. Theory shows that if the Bayes Factor (*BF*_10_) equals 5, this indicates that the data are five times more likely under Hypothesis 1 (*H*_1_) than under Hypothesis 0 (*H*_0_). This means that *H*_1_ provides better probabilistic prediction for the observed data than does *H*_0_ [34]. The Bayesian framework requires the explicit specification of (at least) two models to compare, whereas the frequentist framework relies on only one [37]. Clinical researchers are often interested in several questions when developing a treatment: does the treatment work better, worse, or no differently than an existing placebo or active control? This approach to model comparison through a Bayesian framework allows for a more direct way of answering such questions.

In contrast to p-values, BFs retain their meaning in situations where data are provided over time, regardless of any sampling decisions, therefore data can be analysed repeatedly as it becomes available, without needing special corrections (see Schönbrodt and colleagues [28]). Therefore, the Bayesian approach allows interim analyses to guide decision-making during RCTs, making them attractive when using adaptive trial designs [38, 39].

For this study, BFs were computed repeatedly during interim analyses, starting when 20 participants had been enrolled and approximately every 4-10 participants thereafter, up to a maximum of 150. Early stopping of the trial for either futility or sufficient evidence of benefit was considered if the respective BFs exceeded a pre-defined threshold of 20 which would suggest strong evidence [35]. This sequential BF stopping rule is a suggestion, not a prescription [28]; in Bayesian analyses we are able to sample until hypotheses have been convincingly proven/disproven, or until resources run out [40].

We first calculated a BF for a negative effect of the intervention (i.e. whether participants in the immediate arm had a greater number of IMs at week 4 than the delayed arm). If this BF exceeded 20, we concluded that there was strong evidence for a negative effect of the intervention and the trial may need to be altered or stopped. We then calculated a BF for positive treatment effect (i.e. whether those in the immediate arm had fewer IMs at week 4 than the delayed arm, as opposed to having no difference). If this BF exceeded 20, we concluded that there was strong evidence for the effectiveness of the intervention and consideration could be given to stopping the trial early (Supplementary Information).

Originally, we planned to explore potential ‘mechanistic’ optimisations to improve the effectiveness of the intervention (e.g., time playing Tetris^®^). Given the rapid accumulation of evidence in favour of a positive treatment effect (see ‘Results’), we focused on practical ‘usability enhancements’ to aid smooth digital delivery and user experience (Supplementary Information): this included repeating the intrusive memory visualisation step, adding a summary instruction video, and adding a graphical representation of daily IMs for the four weeks. An optimisation enhancement round was conducted on Feb 7, 2022, after 55 participants had been randomised. When testing for the effect of these enhancements, sample size analyses were first conducted to estimate the number of participants needed to test for a positive treatment effect, and to compare pre-and post-optimisation groups (Supplementary Fig. 8 and 9).

## Data availability

Anonymised databases with the individual participant data and the metadata for Bayesian analyses, along with a data dictionary and analytical code have been uploaded to the Open Science Framework (OSF) and will be made available following publication (< *OSF link to be added here, available during review process*>) for anyone who wishes to access the data for any purpose. The Study Protocol and Bayesian Statistical Analysis Plan are available on the OSF platform (osf.io/2xn5m). All data and supporting information mentioned above will be shared indefinitely and with no end date on the OSF Platform.

## RESULTS

Between Aug 16, 2021 and Apr 19, 2022, 125 participants were screened by the study team following pre-screening questionnaires (Fig. 1). 102 eligible participants provided informed consent and commenced their baseline week diary. Seven participants were then excluded for having fewer than three IMs during baseline week, seven were lost to follow-up, and two participants withdrew prior to randomisation. 86 participants were randomly assigned to the to the delayed (*n*=43) or immediate (*n*=43) arm. Four weeks after randomisation, primary outcome data were available for 75 (87.2%) of randomised participants (Fig. 1), who were taken as our intention-to-treat population [41].

### Baseline characteristics

Trial participants had a mean age of 38.7 years (SD 9.9), were predominantly women (*n*=69; 80.2%) and working full time (*n*=66; 76.7%). The number of IMs experienced in the baseline week were similar between trial arms (Fig. 2A) (combined median=14, IQR=9-20). Baseline characteristics (Supplementary Table 1) appear balanced between arms.

### Treatment effects

Bayesian analyses of the primary outcome involved seven sequential analyses (after 20, 23, 29, 37, 41, 45, and 75 participants completed the primary outcome). From the first analysis (at *n*=20) there was strong evidence against a negative treatment effect (BF=59.8) (Supplementary Table 4). Thereafter, strong evidence in favour of the hypothesis that there was a positive treatment effect rapidly accumulated (Fig. 3). Supportive evidence for the positive treatment effect was reached well before the originally proposed sample size had been randomised (Supplementary Table 4). Following DMC recommendation to the trial steering committee, the trial was concluded early as there was sufficient evidence for the effectiveness of the intervention.

For the final sample (*n*=75), median number of IMs of traumatic events over the seven-day period at week 4 was 1 (IQR=0-3) in the immediate arm, compared with a median of 10 (IQR=6-16.5) in the delayed arm (Fig. 2B), with an estimated Cohen’s *d* effect size of 0.85 (95% CI 0.36 to 1.33). In the fitted Bayesian model, the categorical treatment assignment parameter, which provides a comparison of the immediate arm to the delayed arm (taken as reference level), has posterior mean -1.9 (95% credible interval -2.49, -1.37). This result indicates that when controlling for baseline number of IMs, we would expect the logged number of IMs to be 1.9 lower for those in the immediate arm than the delayed arm.

Equivalently, on the unlogged scale, those in the immediate arm tend to have 0.15 times as many IMs at week 4 as those in the delayed arm.

For the round of ‘usability enhancement’ optimisations (see ‘Method’) conducted on Feb 7, 2022, there was evidence for a positive treatment effect of the optimised intervention (BF=7.31) based on analyses of 28 participants who entered the trial under the optimised intervention.

Sensitivity analyses were completed for variation in Bayesian priors or model used, and to assess impact of outliers and missing data imputation (Supplementary Fig. 4-7). There was no considerable variation in the posterior distributions of our model parameters when varying the Bayesian prior or model used. Outliers were identified as observations with large residuals and large Cook’s distance and leverage; analyses excluding these outliers led to the same pattern of results. In the final analyses, only two participants (one on immediate arm, and one on delayed arm) had one value in their week 4 daily IM diary imputed; analyses excluding these participants also led to similar results (Supplementary Information and Supplementary Fig. 7).

### Safety

By using sequential analyses, we could early (at *n*=20) assure that there was strong evidence against a negative treatment effect (BF=59.8). All adverse and serious adverse events were unrelated to the study (see Supplementary Table 5 and 6): There were 19 adverse events (in 14 participants) in the delayed arm, and 13 adverse events (in 11 participants) and a single serious adverse event (admitted to hospital for chest infection with reduced foetal movement) in the immediate arm.

## DISCUSSION

Results showed strong evidence that ICU staff experiencing IMs after work-related traumatic events in the COVID-19 pandemic, had fewer IMs (median=1 per week, IQR=0-3) when they were given access to the brief digital imagery-competing task intervention, as opposed to usual care for four weeks (median=10 per week, IQR=6-16.5) (Fig. 2). Sequential Bayesian analyses allowed us to rule out any negative effects early in the trial (by *n*=20). Subsequently we were able to conclude the study early – cutting our maximum proposed sample size (*n*=150) substantially. Further, we implemented and assessed intervention enhancements in the same trial, providing evidence that a positive treatment effect was still present after changes. To our knowledge, this is the first RCT to optimise an intervention for the treatment of IMs after traumatic events, and one of the first in mental health to use an adaptive Bayesian approach [42–45].

At trial entry, participants reported a very high number of work-related traumatic events during the pandemic (on average more than 35 traumas, Supplementary Table 1), most of which had taken place over three months ago. Many participants had ongoing trauma exposure during the trial. Prior to the intervention, participants experienced a high number of IMs in daily life – median of 14 per week (baseline, Fig. 2 and Supplementary Table 2), reflecting the symptom burden faced by healthcare workers. In the immediate arm, IMs reduced to a median of one per week, with an average 78% reduction in the number of IMs compared to baseline (Supplementary Table 2), and 36% experiencing zero IMs at week 4 (Supplementary Fig. 1).

There remains an urgent need for scalable approaches to support the mental health of frontline healthcare workers. Given their high workload demands, we developed a brief and flexible digital imagery-competing task intervention approach to reduce IMs. After one initial session with research guidance, the intervention could thereafter be used independently and was repeatable to treat different IMs and new trauma (e.g. intrusive image of a dying patients face; intrusive image of colleague on ambulance stretcher; etc). Compared to studies on the day of trauma [8, 14], current results offer the possibility to deliver treatment when a longer time has elapsed (i.e., weeks or months post-trauma), and thus be useful for individuals already experiencing IMs such as ICU staff.

Adopting new statistical approaches for RCTs is an essential step in speeding up the development of new interventions, and associated moral and ethical decisions in the use of RCTs. By utilising advances in Bayesian trial methodology in the present study to optimise the brief digital imagery-competing task intervention, we substantially reduced the sample size and therefore the time and resources needed to run the trial (Fig. 3). This allowed a more efficient trial without sacrificing statistical and/or scientific rigour. By the explicit model comparison aspect of the Bayesian approach and ability to quantify evidence for the hypotheses of interest using BFs, we could determine, more rapidly, whether the intervention provided a negative or positive effect on the frequency of IMs. The fact that BFs retain their meaning in situations where data is collected over time, allowed us to monitor evidence in an almost continuous manner using sequential analyses, affording the opportunity to exploit the advantages of adaptive trial designs. Based on recent studies on the relationship between p-values and BFs, our results are arguably even more robust than if we had merely determined a frequentist p-value to some level of significance [26].

In general, the practical consequences of using more efficient adaptive Bayesian trial designs to develop therapeutic approaches are clear – they can provide information rapidly to support a go/no-go clinical development decision, thus helping treatment innovation by reducing the time required to progress to subsequently assess efficacy. Here, it helped us examine the effects of a new intervention approach. Methods advances are needed given the relatively slow progress of behavioral interventions since the 1960s [47, 48]. This Bayesian study focuses on the primary outcome –a fuller set of analyses using frequentist statistical approaches will be reported in a companion article [33].

In retrospect, we could have conducted an even more efficient trial. As the evidence progression shows, there was sufficient evidence (BF>20) when 29 participants had completed the trial to conclude that there were positive effects of the intervention. However, at that point to ensure the robustness of the results we conducted a more thorough sensitivity analyses. Simultaneously, we strived for equivalent allocation to the two arms (immediate and delayed), a balancing that ultimately delayed the conclusion of the study. Such sensitivity and balancing issues can be mitigated beforehand [49]. Further limitations of this trial include the use of a wait list design. The design was chosen due to the early stage of the intervention development, the lack of a comparator treatment for IMs, and ethical considerations around the participant population. The statistician running Bayesian analyses (VR) was not blinded to group allocation and was a part of the wider study team. There is a reduction of IMs in both arms, and placebo effects cannot be ruled out. The next study should use a comparison arm rather than wait list.

There is an urgent and unmet need to develop novel approaches to support the mental health of healthcare workers to continue to manage the emotionally traumatic nature of their clinical work [7]. Current clinical guidelines show we lack treatment approaches for people facing ongoing trauma exposure, such as those working in the ICU (Supplementary Table 1) [50].

Addressing the mental health challenges of healthcare workers is important for them as individuals but is also important for the sustainability of the provision of healthcare services, particularly during a pandemic and in the recovery of health services post-pandemic. In this study, we addressed the need for accelerating treatment development by the use of Bayesian methodology, essentially cutting sample size in half while allowing for testing more hypotheses than in a traditional frequentist trial. Results showed strong evidence in favour of a positive treatment effect of the brief (one guided-session), remotely delivered digital intervention in reducing the number of IMs after trauma. Next steps include a trial with a control comparator. Overall, for ICU staff with unwanted intrusive images of traumatic events from work, this optimisation trial during the pandemic suggests that a brief digital imagery-competing task intervention may help reduce the frequency at which trauma memories intrude. This novel intervention shows promise for further development.

## Supporting information

Supplementary Information (including Supplementary Figures and Tables)

## Data Availability

Anonymised databases with the individual participant data and the metadata for Bayesian analyses, along with a data dictionary and analytical code have been uploaded to the Open Science Framework (OSF) and will be made available following publication for anyone who wishes to access the data for any purpose. The Study Protocol and Bayesian Statistical Analysis Plan are available on the OSF platform (osf.io/2xn5m). All data and supporting information mentioned above will be shared indefinitely and with no end date on the OSF Platform.

## ACKNOWLEDGMENTS

This work was supported by the Wellcome Trust (223016/Z/21/Z)

For the purpose of open access, the author has applied a CC BY public copyright licence to any Author Accepted Manuscript version arising from this submission.

Tetris^®^ has been licenced for use within i-spero^®^ from The Tetris Company.

The authors would like to thank the Intensive Care Society, in particular Dr Sandy Mather and Alex Day; Our Data Monitoring Committee members including Prof Andreas Reif, Prof Steve Hollon, and Prof Ian Penton-Voak; Trial Steering Committee including Prof Guy Goodwin, Pooyan Behbahani, and Rebecca Dias; Expert Advisors including Dr Nick Grey, Prof Sir Simon Wessely, and Prof Jonathon Bisson. Members of study team including Veronika Kubičková, Alfred Markham, Zunaid Islam and Marie Kanstrup for support; Our Data Management Team including Mark Dziedzic, Sameer Iqbal, and Nikita Shukan.

## CONFLICT OF INTEREST

The study was funded by the Wellcome Trust (223016/Z/21/Z). JK is shareholder and director of P1vital Products Ltd which is the study sponsor and manufacturer of i-spero^®^. VR and LI are employed by P1vital Products Ltd. MBB is an adjunct member of the DMC. CS salary is part funded by National Institute for Health Research (NIHR) and Medical Research Council (MRC). EAH is on the Board of Trustees of the MQ Foundation. EAH also receives funding from the Swedish Research Council, Rannís The Icelandic Research Fund, OAK foundation, The Lupina Foundation and AFA Försäkring. EAH developed the intervention approach and training in using it (Anenome ™). EAH receives book royalties from Guildford Press and Oxford University Press, and receives occasional honoraria for conference keynotes and clinical workshops. All other authors declare no competing interests.

## AUTHOR CONTRIBUTIONS

EAH and JK conceived the trial, gained funding, and contributed to the study design. CS and MBB provided expert input into the conception of the trial, and MBB and TJ into the study design. LI delivered the intervention, acquired the data, and with JK, JH and CS contributed to interpretation of the work. VR and MBB accessed and verified the underlying data. VR did the statistical analyses, and with MBB, EAH and PM contributed to the analysis of the work and interpretation of data. VR, PM, EAH, and MBB wrote the first draft of the article. All authors contributed intellectual content, critically revised the article, and approved the submitted manuscript.

## FUNDING

The study was funded by the The Wellcome Trust (223016/Z/21/Z). The funder of the study had no role in study design, data collection, data analysis, data interpretation, or writing of the report. P1vital Products Ltd was the trial sponsor.

## SUPPLEMENTARY INFORMATION

Supplementary Information (including Supplementary Table 1 through 6 and Fig. 1 through 9) is provided as a separate combined pdf file.

## FIGURES

Fig. 1 through 3 are provided below in numerical order.

## Notes

### Clinical Trial

NCT04992390

### Clinical Protocols

https://osf.io/2xn5m/

### Funding Statement

This research was funded in whole or in part by the Wellcome Trust (223016/Z/21/Z). 

### Author Declarations

Wales Research Ethics Committee (Wales REC 6, 21/WA/0173) of the NHS Health Research Authority gave ethical approval for this work

### Summary of Updates

The clinical trial registration (CTR) link in very last sentence of the abstract was corrected to: Trial Registration NCT04992390. We have removed our own header and footer from the manuscript title page as this included the same details as the header and footer added by medRxiv.

