## Supplementary Information (including Supplementary Figures and Tables) for "Treating intrusive memories after trauma in healthcare workers: a Bayesian adaptive randomised trial developing an imagery-competing task intervention"

1. Statistical Analyses
   1.1. Software
   1.2. Model Framework
   1.3. Missing Data
   1.4. Model Fit
   1.5. Bayes Factors
   1.6. Sensitivity Analyses
      1.6.1. Priors
      1.6.2. Model Choice
      1.6.3. Outliers
      1.6.4. Missing Data Imputation
2. Optimisations to the Intervention
   2.1. List of Optimisations
   2.2. Sample Size Calculation
   2.3. Evidence for Positive Treatment Effect
3. Results from Interim Analysis
4. References for Supplementary Information
5. Table Legends
6. Tables
7. Figure Legends
8. Figures

**NOTE: This preprint reports new research that has not been certified by peer review and should not be used to guide clinical practice**

### 1. STATISTICAL ANALAYSES

#### 1.1. Software

All analyses were conducted using R version 4.1.2 with recent software versions available at the time [1]. The R package *tidyverse* was used for data manipulation and plotting with *ggplot2*. Bayesian models were fitted using the R package *brms* [2, 3]. Model fit was evaluated through built-in posterior checks in *brms*. The package *bayestestR* was used to calculate Bayes Factors (BFs) for the hypotheses relating to the primary objective. Exploratory modelling and model comparisons were conducted using R packages: *lme4, MASS, pscl, boot, performance, ZIM,* and *lmtest*. Summary statistics were created using the *table1* package in R.  Bespoke R scripts using time series models implemented in an E-M approach were used to deal with imputation of missing values (for details on the algorithm please refer to '1.3 Missing Data') [4].

#### 1.2. Model Framework

The primary outcome is based on count data. Therefore, a Bayesian Poisson generalised linear mixed model was used to examine group difference in the primary outcome [5–8], with the baseline number of intrusive memories (at run-in week), and treatment assignment taken as fixed effects. Let $n$ be the number of participants to be analysed, then for each participant $i = 1, \dots, n$ we denote:

- $Y_i$ to be the random variable representing the primary endpoint (total number of intrusive memories recorded during week 4)
- $Baseline_i$ to be the baseline number of intrusive memories (total number of intrusive memories recorded during the run-in week)

- $ARM_i$ a categorical variable with two levels that represents whether the participant is in the immediate intervention arm or in the delayed intervention arm (control arm, taken as the reference factor level)

Exploratory data analysis was first used to investigate the distribution of the primary outcome measure. Through visual and quantitative analysis, the validity of any model assumptions were examined. Poisson, Poisson with random intercept effect for participant (Observation-level random effects Poisson model) [9], Negative Binomial, Zero Inflated Poisson, and Zero Inflated Negative Binomial general linear models were compared using various tests and performance measures such as Likelihood Ratio Tests, AIC and BIC, Root Mean Squared error and BFs. The most appropriate model which corrected for any zero inflation and overdispersion present was used.

A normal prior with (mean 0, standard deviation 10) was taken for the population level effects of the intervention arm and baseline number of intrusive memories. The default priors set within the ***brms*** package [2, 3] were used for any other model parameters such as the intercept, any group-level effects, or any family specific parameters. A student-t prior (with 3 degrees of freedom, location parameter of 1.1, and scale parameter of 2.5) was used for the intercept parameter $\alpha$. The standard deviation parameter for the random effect was restricted to be positive and takes a half student-t prior (with 3 degrees of freedom and a scale parameter 2.5). Negative Binomial and Zero Inflated Negative Binomial need a shape parameter that had a gamma (shape 0.01, scale 0.01) prior by default. To transform the linear predictor of any zero inflated parameters into a probability, *brms* applies the logit-link, which takes values within [0.00, 1.00] interval and returns values on the real line. Thus, it allows the transition between probabilities and linear predictors.

The observation model used for the final analyses was Poisson with random intercept effect for participant using priors as described below. The model is as follows where $\alpha, \beta_1, \beta_2$ are the fixed population level effects and $\gamma_{0i}$ are random effects whose variances are estimated by the model. $\gamma_{0i}$ is the random effect for the intercept for each participant $i$ that accounts for the participant-specific variation in the primary endpoint. The random-effects intercepts $\gamma_{0i}$ are drawn from a normal distribution with mean 0 and variance $\sigma_{\gamma_0}$ which is estimated by the model.

$$Y_i \sim Poisson(\mu_i)$$

$$log(\mu_i) = \alpha_i + ARM_i\beta_1 + Baseline_i\beta_2$$

$$\alpha_i = \alpha + \gamma_{0i}$$

$$\gamma_{0i} \sim Normal(0, \sigma_{\gamma_0})$$

##### 1.3. Missing Data

When genuine missing data occurred, we imputed the missing values using time-series methods [10] and an expectation-maximisation (E-M) algorithm [4]. Initial missing values were imputed by taking expectations across a participant's available diary data. Using Poisson likelihood and correlated errors, we maximised over this 'full' data set to provide updated expected values for the missing data. We iterated over these latter steps until convergence in values of missing data (to a pre-determined threshold) was achieved.

Let $y = (y_1, y_2, y_3, y_4, y_5, y_6, y_7)$ be a participant's observed diary data. The maximum likelihood approach for dealing with missing values fits an autoregressive model of order 1 (AR(1) model) regression to the observed data through a simulated annealing algorithm. The main algorithm uses an E-M algorithm to impute missing values based on Poisson time series regression [4]. Given a linear time-series model $y_t = a_1 y_{t-1} + a_2$, the mean of a Poisson distribution is $\lambda = exp(a_1 y_{t-1} + a_2)$, with Poisson probability function

$$Pr(y_t \mid y_{t-1};\ a_1, a_2) = \frac{\lambda^{y_t}}{y_t!} exp(-\lambda).$$

Across the time series, the conditional likelihood is $L(a_1, a_2 \mid y_t, y_{t+1}) = \prod_{i=2}^{7} Pr(y_i \mid y_{i-1};\ a_1, a_2)$.

Initial imputation uses random draws from a Poisson with mean $\bar{y}$ to impute the missing values. The E-M algorithm then maximizes over this ‘full’ data set and updates the missing values based on model predictions.

We saw only a few participants having genuine missing data in their intrusive memory diaries during run-in week and week 4. In the final analyses, two participants (one on immediate arm, and one on delayed arm) had one value in their week 4 daily IM diary imputed using the method detailed above. See ‘1.6 Sensitivity Analyses’ for details of sensitivity analyses undertaken for missing data imputation.

##### 1.4. Model Fit

The Bayesian models were fitted using 4 chains, each with 20,000 iterations of which the first 1000 were warmup to calibrate the sampler, leading to a total of 76,000 posterior samples. Graphical diagnostic plots made using the *plot* function within the *brms* package were inspected to check for convergence. From Supplementary Fig. 2, we see that the Markov chain Monte Carlo (MCMC) algorithm converged and mixes well for all parameters in the model. Posterior predictive checks were performed using the *pp_check* function in the *brms* package to check whether the distributional assumption of the model is reasonable (see Supplementary Fig. 3).

A summary of the fitted Bayesian model for the final analyses is provided in Supplementary Table 3. This coefficient table is based on the posterior distributions of the estimated parameters, the principal outcome of fitting a Bayesian model. The posterior distribution is a probability distribution that indicates how probable particular parameter values are, given the prior distribution and the observed data. In the Bayesian model, the 95% credibility interval states that there is 95% chance that the true population value falls within this interval. We see that the 95% credibility interval for the Intervention Arm parameter falls below zero, so we conclude that this model parameter is likely meaningful and that the model predicts fewer intrusive memories for those on the immediate intervention arm than for those on the delayed intervention arm [2, 3].

##### 1.5. Bayes Factors

The BF is either denoted as $BF_{01}$ ("$H_0$ over $H_1$,") or as its inverse $BF_{10}$ ("$H_1$ over $H_0$"). When the $BF_{01}$ equals 5, this indicates that the data are five times more likely under $H_0$ than under $H_1$, meaning that $H_0$ has issued a better probabilistic prediction for the observed data than did $H_1$ [11].

First, we tested for evidence against no benefit (a negative effect) of the intervention:

| | |
|---|---|
| $H_1$: $\beta_1 > 0$ | The intervention has a negative effect (participants in the immediate intervention arm, compared to the delayed intervention arm, will have a greater number of intrusive memories in week 4) |
| $H_0$: $\beta_1 \leq 0$ | The intervention has no effect or a positive effect (participants in the immediate intervention arm, compared to the delayed intervention arm, will have equal or fewer intrusive memories) |

If $BF_{10}$ exceeded 20 for the above hypotheses (equivalently if $BF_{01}$ is less than $\frac{1}{20}$), we concluded that there was strong evidence for no benefit (a negative effect) of the intervention (i.e. that those in the immediate intervention arm, compared to the delayed intervention arm, have a greater number of intrusive memories) and the trial may need to be altered or stopped.

Second, testing for positive treatment effect of the intervention:

| | |
|---|---|
| $H_1: \beta_1 < 0$ | The intervention has a positive effect (participants in the immediate intervention arm, compared to the delayed intervention arm, will have fewer intrusive memories) |
| $H_0: \beta_1 = 0$ | The intervention has no effect (participants in the immediate intervention arm, compared to the delayed intervention arm, will have an equal number of intrusive memories in week 4) |

If $BF_{10}$ exceeded 20 for the above hypotheses (equivalently if $BF_{01}$ is less than $\frac{1}{20}$), we concluded that there was strong evidence for the effectiveness of the intervention (i.e. that those in the immediate intervention arm, compared to delayed intervention arm, have fewer intrusive memories) to consider concluding the trial early. Data collection could be stopped whenever the evidential threshold has been exceeded, or when the a priori defined maximal sample size of 150 has been reached.

#### 1.6. Sensitivity Analyses

##### 1.6.1. Priors

We examined how robust the results are when the priors taken for the population level effects of the intervention arm and baseline number of intrusive memories were altered, and the model was re-estimated. This included comparing posterior distribution of the relevant model parameters.

Our results, displayed in Supplementary Fig. 4, show that the posterior distributions of the relevant model parameters do not vary significantly when tested with different prior distributions.

##### 1.6.2. Model Choice

We examined how robust the results were to the choice of model used (Poisson, Poisson with random intercept effect for participant, Negative Binomial, Zero Inflated Poisson, and Zero Inflated Negative Binomial) to model the primary outcome. This included comparing posterior distribution of the relevant model parameters.

Our results, displayed in Supplementary Fig. 5 show that the posterior distributions of the relevant model parameters vary only by a small amount when different models were used; we see that the 95% credible interval for the fixed population level effect of the treatment arm are negative across all models lying in the interval [-2.49, -0.40]. The 95% credible interval stays around 0 for the fixed population effect of the baseline number of intrusions across all the models, lying in the interval [0.00, 0.08].

##### 1.6.3. Outliers

Analysis was conducted using all data available. Outliers were identified through inspection of residual plots and Cook's distance vs leverage plots under a Poisson model. Where outliers were identified, results using data without outliers were examined as sensitivity analysis: this included comparing posterior distributions of the relevant model parameters, and the BFs when the model was re-estimated.

In the final analyses, we identified three outliers. With these outliers excluded and the model re-estimated, the posterior distributions of the relevant model parameters do not vary significantly (see Supplementary Fig. 6). Excluding these outliers did not affect the conclusion that there was strong positive treatment effect (strong evidence = BF > 20, BF when including outliers = $1.25 \times 10^{6}$, BF when excluding outliers = $7.77 \times 10^{7}$).

###### 1.6.4. Missing Data Imputation

Analysis was conducted using all data available. In the final analyses, two participants (one on immediate arm, and one on delayed arm) had one value in their week 4 diary imputed used the method detailed in 'Missing Data'. Results excluding these participants were examined as sensitivity analysis: this included comparing posterior distributions of the relevant model parameters, and the BFs when the model was re-estimated.

With the participants who had imputed values excluded and the model re-estimated, the posterior distributions of the relevant model parameters do not vary significantly (see Supplementary Fig. 7). Excluding these participants did not affect the conclusion that there was strong positive treatment effect (strong evidence = BF>20, BF for all data = $1.25 \times 10^{6}$, BF when using non-imputed data only = $3.03 \times 10^{7}$).

#### 2. OPTIMISATIONS TO THE INTERVENTION

##### 2.1. List of Optimisations

Details of the 'usability enhancement' optimisations which were implemented on Feb 7, 2022:

i. Repeat intrusive memory visualisation immediately before playing Tetris®

ii. Additional informational video on how to use the intervention in daily life

iii. Prevent intervention being completed after user records their daily intrusive memories.

iv. Allow intervention feedback questionnaire to be completed from 35 to 42 days after the first guided intervention session.

**v.** Additional visual feedback of recorded intrusive memory results over time.

##### 2.2. Sample Size Calculation

We conducted sample size analyses to guide recruitment and determine which tests we can conduct to assess the optimised intervention using the R package *bayesmedr*. The sample size analyses (visualised in Supplementary Fig. 8) illustrated that we could assess the optimised intervention by testing for a positive treatment effect. Using the graphs we estimated that we must recruit around 20 participants in total under the enhancement optimisations to see strong evidence (BF of 20) in favour of a positive treatment effect compared to no effect (assuming roughly equal arm split, and similar primary outcome means as seen in data before optimisation). In Supplementary Fig. 9 we see that the BFs do not reach 20 for the tested sample size ranges, we therefore concluded that it was not feasible within the project timeline to conduct a comparison between the optimised intervention and the former un-optimised intervention.

##### 2.3. Evidence for Positive Treatment Effect

We tested for positive treatment effect $(H_1)$ against no effect $(H_0)$ of the intervention by analysing only the participants who entered the trial under the optimised intervention. We found a BF of 7.31 in favor of $H_1$, meaning there was moderate evidence that the optimised version of the intervention also resulted in a positive treatment effect (based on analyses of 28 participants who entered the trial under the optimised intervention).

#### 3. RESULTS FROM INTERIM ANALYSIS

In Supplementary Table 4, we present the BFs calculated for the two sets of hypotheses detailed in Section '1.5 Bayes Factors' using data from the Nov 24, 2021 and onwards. We see that strong evidence against no benefit (a negative effect) of the intervention was present from the first interim analyses using data from Nov 24, 2021 and onwards. Strong evidence in favour of positive treatment effect was present using data from Dec 22, 2021 and onwards.

7. Iyadurai L, Blackwell SE, Meiser-Stedman R, Watson PC, Bonsall MB, Geddes JR, et al. Preventing intrusive memories after trauma via a brief intervention involving Tetris

computer game play in the emergency department: A proof-of-concept randomized controlled trial. Mol Psychiatry. 2018;23:674–682.

8. James EL, Bonsall MB, Hoppitt L, Tunbridge EM, Geddes JR, Milton AL, et al. Computer Game Play Reduces Intrusive Memories of Experimental Trauma via Reconsolidation-Update Mechanisms. Psychol Sci. 2015;26:1201–1215.

9. Harrison XA. Using observation-level random effects to model overdispersion in count data in ecology and evolution. PeerJ Inc. 2014;2.

10. Chatfield C and AJCollins, & BAF. The analysis of time series: an introduction. Sixth Edition. New York: Chapman and hall/CRC; 2003.

11. Schönbrodt FD, Wagenmakers EJ. Bayes factor design analysis: Planning for compelling evidence. Psychon Bull Rev. 2018;25:128–142.

#### 5. TABLE LEGENDS

**Supplementary Table 1. Baseline Characteristics and Number of Traumatic Events Data Summaries.** Demographics, Work and Employment, Health Background, Experiences of Prior Trauma, and Experiences of Ongoing Trauma are presented for all randomised participants by arm (delayed arm (control), n = 43: usual care for four weeks; immediate arm, n = 43: immediate access to the intervention following the baseline week). Mean (SD) for continuous variables and n (%) for categorical variables are presented. The continuous variables for 'Experiences of Ongoing Trauma' have been categorised into 5 groups.

**Supplementary Table 2. Number of Intrusive Memories Data Summaries.** Minimum (Min) and Maximum (Max), Median, Upper and Lower Quartiles, Mean, Standard deviation (SD) are presented by arm (delayed arm (control), n = 39: usual care for four weeks; immediate arm, n = 36: immediate access to the intervention following the baseline week), and in total for all participants included in final interim Bayesian analyses (n=75). Data summaries are included for: the total number of intrusive memories recorded over 7 days during the run-in/baseline week, the total number of intrusive memories recorded over 7 days during week 4 (primary outcome), and the ratio reduction in the total number of intrusive memories recorded between run-in/baseline week to week 4

$$\left(\frac{\text{no. of intrusive memories at baseline} - \text{no. of intrusive memories at week 4}}{\text{no. of intrusive memories at baseline}},\ \text{no. of intrusive memories at baseline} > \mathbf{0}\right)\textbf{.}$$

Summary statistics created using *table1* package in R with default settings.

**Supplementary Table 3. Model Summary.** For fitted Bayesian model in final analyses, every model parameter is summarised using the mean (Estimate) and the standard deviation (Estimated Error) of the posterior distribution as well as two-sided 95% credible intervals (Lower 95% Credible Interval and Upper 95% Credible Interval) based on quantiles. Tail Effective Sample Size (Tail-ESS) is obtained by computing the minimum of effective sample sizes for 5% and 95% quantiles. Bulk ESS; is the number of independent samples from the posterior distribution that would be expected to yield the same standard error of the posterior mean as is obtained from the dependent samples returned by the MCMC algorithm. The Rhat value provides information on the convergence of the algorithm: The Rhat value are all near 1, suggesting that the chains have converged and mixed well.

**Supplementary Table 4. Bayes Factors.** Bayes Factors calculated using the data transfers from the Nov 24, 2021 onwards. Table columns show the date of data transfer, the number of participants analysed, and the Bayes Factor which test the two hypotheses as described in Section ‘Bayes Factors’. The cells colour coded in green indicate when the a priori defined threshold to consider early stopping of the trial was passed.

**Supplementary Table 5. Serious adverse events.** Serious adverse events categories observed in the delayed and the immediate intervention arms. Only one serious adverse event was self-reported by a participant in the immediate arm. All serious adverse events were unrelated to the study.

**Supplementary Table 6: Adverse events.** Adverse event categories observed in the delayed and the immediate intervention arms. NOS = not otherwise specified. In the immediate arm 13 adverse events (in 11 participants) were recorded, and in the delayed arm 19 adverse events (in 14 participants) were recorded. Adverse events were reported by participants either when completing secondary outcome questionnaires at baseline, week 4 and week 8, or were self-reported during contact with researchers. All adverse events were unrelated to the study.

#### 6. TABLES

Supplementary Tables 1 through 6 are provided below in numerical order.

| Demographics | Delayed arm (n=43) | | Immediate arm (n=43) | |
|---|---|---|---|---|
| | **Mean** | **SD** | **Mean** | **SD** |
| Age (years) | 39.9 | 9.9 | 37.4 | 9.8 |
| | **n** | **%** | **n** | **%** |
| Gender | | | | |
| Woman | 35 | 81.4% | 34 | 79.1% |
| Man | 7 | 16.3% | 8 | 18.6% |
| Transwoman | 0 | 0 | 0 | 0 |
| Transman | 0 | 0 | 0 | 0 |
| Gender-variant/non-binary | 0 | 0 | 0 | 0 |
| Other Identity | 0 | 0 | 0 | 0 |
| Prefer not to answer | 0 | 0 | 0 | 0 |
| Missing | 1 | 2.3% | 1 | 2.3% |
| Highest level of education | | | | |
| Secondary school (to age 16) | 1 | 2.3% | 0 | 0 |
| Sixth form or equivalent (to age 18) | 1 | 2.3% | 3 | 7.0% |
| Bachelor's degree or equivalent | 23 | 53.5% | 24 | 55.8% |
| Master's degree | 13 | 30.2% | 11 | 25.6% |
| Doctoral degree | 3 | 7.0% | 3 | 7.0% |
| Prefer not to answer | 1 | 2.3% | 1 | 2.3% |
| Missing | 1 | 2.3% | 1 | 2.3% |
| Ethnicity | | | | |
| White - British | 18 | 41.9% | 18 | 41.9% |
| White - Irish | 1 | 2.3% | 1 | 2.3% |
| White - Any other White background | 14 | 32.6% | 9 | 20.9% |
| Mixed - White and Black Caribbean | 0 | 0 | 0 | 0 |
| Mixed - White and Black African | 0 | 0 | 0 | 0 |
| Mixed - White and Asian | 0 | 0 | 0 | 0 |
| Mixed - Any other mixed background | 0 | 0 | 2 | 4.7% |
| Asian - Indian | 3 | 7.0% | 2 | 4.7% |
| Asian - Pakistani | 0 | 0 | 0 | 0 |
| Asian - Bangladeshi | 0 | 0 | 0 | 0 |
| Asian - Any other Asian background | 0 | 0 | 1 | 2.3% |
| Black - Caribbean | 0 | 0 | 0 | 0 |

| | | | | |
|---|---|---|---|---|
| Black - African | 1 | 2.3% | 1 | 2.3% |
| Black - Any other Black background | 0 | 0 | 0 | 0 |
| Other - Chinese | 0 | 0 | 0 | 0 |
| Other - Any other ethnic group | 1 | 2.3% | 0 | 0 |
| Other - Traveller | 0 | 0 | 0 | 0 |
| Other - Arab | 0 | 0 | 0 | 0 |
| Unknown - Not stated | 4 | 9.3% | 8 | 18.6% |
| Missing | 1 | 2.3% | 1 | 2.3% |
| Marital status | | | | |
| Single | 13 | 30.2% | 13 | 30.2% |
| Living apart from partner | 0 | 0 | 1 | 2.3% |
| Married or cohabiting | 28 | 65.1% | 25 | 58.1% |
| Divorced or separated | 1 | 2.3% | 3 | 7.0% |
| Widowed | 0 | 0 | 0 | 0 |
| Other | 0 | 0 | 0 | 0 |
| Missing | 1 | 2.3% | 1 | 2.3% |
| **Work and Employment** | | | | |
| | **Mean** | **SD** | **Mean** | **SD** |
| Workplace | | | | |
| Work time (hours per week) | 37.8 | 9.2 | 37.9 | 13.3 |
| Time as healthcare professional (years) | 16.4 | 10.8 | 13.0 | 8.6 |
| | **n** | **%** | **n** | **%** |
| Employment status | | | | |
| Working full time | 32 | 74.4% | 34 | 79.1% |
| Working part time | 10 | 23.3% | 6 | 14.0% |
| Jobseeking | 0 | 0 | 0 | 0 |
| Sick leave | 0 | 0 | 1 | 2.3% |
| Student | 0 | 0 | 0 | 0 |
| Retired | 0 | 0 | 0 | 0 |
| Other | 0 | 0 | 1 | 2.3% |
| Missing | 1 | 2.3% | 1 | 2.3% |
| NHS Job Role | | | | |
| Allied Health Professionals | 1 | 2.3% | 3 | 7.0% |
| Doctors | 11 | 25.6% | 8 | 18.6% |
| Health Informatics | 1 | 2.3% | 0 | 0 |
| Healthcare Support Worker | 1 | 2.3% | 2 | 4.7% |
| Nursing | 28 | 65.1% | 29 | 67.4% |

| | | | | |
|---|---|---|---|---|
| Other/unknown | 1 | 2.3% | 0 | 0 |
| Pharmacy | 0 | 0 | 1 | 2.3% |
| **Health background** | | | | |
| | **n** | **%** | **n** | **%** |
| Do you have any current physical health problems e.g. diabetes, heart problems? (yes/no, n = yes) | 9 | 20.9% | 8 | 18.6% |
| Have you been treated for/diagnosed with any mental health problems e.g. depression, anxiety, post-traumatic stress disorder? (yes/no, n = yes) | 24 | 55.8% | 22 | 51.2% |
| Has anyone in your close family been treated for/diagnosed with any mental health problems? (yes/no, n = yes) | 16 | 37.2% | 14 | 32.6% |
| Missing | 1 | 2.3% | 2 | 4.7% |
| **Experiences of Prior Trauma** | | | | |
| | **Mean** | **SD** | **Mean** | **SD** |
| How many work-related traumatic events have you experienced/witnessed during COVID-19? Remember: A traumatic event is defined as an event that involved actual or risk of death, serious injury, or sexual violence for you or someone else." | 38.4 | 80.9 | 35.9 | 58.0 |
| How many traumatic events that were not work-related have you experienced/witnessed during COVID-19 (e.g., serious accident, assault, injury or illness)? | 10.2 | 46.2 | 4.5 | 15.6 |
| | **n** | **%** | **n** | **%** |
| Which of the following categories best fit the work-related traumatic events that you have experienced or witnessed during COVID-19, of which you have intrusive memories. | | | | |
| A traumatic or tragic death of a patient | 40 | 93.0% | 38 | 88.4% |
| A severe or unsuccessful resuscitation | 32 | 74.4% | 26 | 60.5% |
| Witnessing events surrounding colleague who has fallen ill or died of COVID-19 | 18 | 41.9% | 19 | 44.2% |
| Situation where the care of a patient failed or did not go as planned | 31 | 72.1% | 33 | 76.7% |
| Threats or violence against healthcare professionals | 13 | 30.2% | 26 | 60.5% |
| Event involving sudden increased risk of COVID-19 infection | 29 | 67.4% | 23 | 53.5% |
| A traumatic or tragic event where a patient reminded you of yourself, a family member or friend | 31 | 72.1% | 29 | 67.4% |

| | | | | |
|---|---|---|---|---|
| Event involving extremely distressed/grieving relatives of patients | 33 | 76.7% | 37 | 86.0% |
| Being faced with suicide / suicide attempt | 18 | 41.9% | 14 | 32.6% |
| Other | 2 | 4.7% | 5 | 11.6% |
| Missing | 1 | 2.3% | 1 | 2.3% |
| Timeframe of traumas experienced at Baseline | | | | |
| Within the last 24 hours | 4 | 9.3% | 3 | 7.0% |
| Within the past month | 8 | 18.6% | 7 | 16.3% |
| Between 1-3 months ago | 7 | 16.3% | 8 | 18.6% |
| More than 3 months ago | 24 | 55.8% | 22 | 51.2% |
| Ongoing exposure to traumatic events is part of my job during the COVID-19 pandemic | 37 | 86.0% | 33 | 76.7% |
| Missing | 1 | 2.3% | 1 | 2.3% |
| | **Mean** | **SD** | **Mean** | **SD** |
| Perceived life threat to someone else | 8.3 | 2.8 | 9.2 | 1.4 |
| Perceived life threat to self | 5.3 | 3.4 | 4.7 | 3.4 |
| Peritraumatic Distress Inventory Total Score | 28.6 | 9.9 | 30.5 | 10.2 |
| **Experiences of Ongoing Trauma** | | | | |
| | **n** | **%** | **n** | **%** |
| ***Week 4*** | | | | |
| Have you experienced or witnessed any new work-related traumatic events? (yes/no, n = yes) | 19 | 44.2% | 11 | 25.6% |
| How many new work-related traumatic events have you experienced/witnessed? | | | | |
| 0 | 16 | 37.2% | 15 | 34.9% |
| 1 - 5 | 19 | 44.2% | 11 | 25.6% |
| 6 - 10 | 1 | 2.3% | 1 | 2.3% |
| 11 - 15 | 0 | 0 | 1 | 2.3% |
| 15+ | 2 | 4.7% | 0 | 0 |
| How many new traumatic events that were not work-related have you experienced/witnessed? | | | | |
| 0 | 32 | 74.4% | 25 | 58.1% |
| 1 - 5 | 6 | 14.0% | 3 | 7.0% |
| 6 - 10 | 0 | 0 | 0 | 0 |
| 11 - 15 | 0 | 0 | 0 | 0 |
| 15+ | 0 | 0 | 0 | 0 |

| | | | | |
|---|---|---|---|---|
| Missing (for all above) | 5 | 11.6% | 15 | 34.9% |
| ***Week 8*** | | | | |
| Have you experienced or witnessed any new work-related traumatic events? (yes/no, n = yes) | 9 | 20.9% | 16 | 37.2% |
| How many new work-related traumatic events have you experienced/witnessed? | | | | |
| 0 | 22 | 51.2% | 14 | 32.6% |
| 1 - 5 | 7 | 16.3% | 13 | 30.2% |
| 6 - 10 | 0 | 0 | 2 | 4.7% |
| 11 - 15 | 2 | 4.7% | 2 | 4.7% |
| 15+ | 0 | 0 | 0 | 0 |
| How many new traumatic events that were not work-related have you experienced/witnessed? | | | | |
| 0 | 28 | 65.1% | 23 | 53.5% |
| 1 - 5 | 3 | 7.0% | 4 | 9.3% |
| 6 - 10 | 0 | 0 | 0 | 0 |
| 11 - 15 | 0 | 0 | 0 | 0 |
| 15+ | 0 | 0 | 0 | 0 |
| Missing (for all above) | 12 | 27.9% | 12 | 27.9% |

| Number of Intrusive Memories (IMs) | All participants (n = 75) | Delayed arm (n = 39) | Immediate arm (n = 36) |
|---|---|---|---|
| **Baseline Week** | | | |
| Min | 3.00 | 5.00 | 3.00 |
| Lower Quartile (25%) | 9.00 | 8.00 | 10.0 |
| Median | 14.0 | 14.0 | 14.5 |
| Mean | 17.8 | 17.1 | 18.5 |
| SD | 14.7 | 15.8 | 13.6 |
| Upper Quartile (75%) | 20.0 | 18.5 | 21.3 |
| Max | 99.0 | 99.0 | 75.0 |
| **Primary Outcome (Week 4)** | | | |
| Min | 0 | 2.00 | 0 |
| Lower Quartile (25%) | 1.00 | 6.00 | 0 |
| Median | 5.00 | 10.0 | 1.00 |
| Mean | 8.41 | 12.5 | 4.03 |
| SD | 10.8 | 9.28 | 10.7 |
| Upper Quartile (75%) | 11.0 | 16.5 | 3.00 |
| Max | 61.0 | 37.0 | 61.0 |
| **Ratio reduction in IMs between Baseline and Week 4** | | | |
| Min | -1.10 | -0.783 | -1.10 |
| Lower Quartile (25%) | 0.120 | -0.107 | 0.764 |
| Median | 0.632 | 0.200 | 0.933 |
| Mean | 0.470 | 0.182 | 0.783 |
| SD | 0.506 | 0.394 | 0.422 |
| Upper Quartile (75%) | 0.929 | 0.509 | 1.00 |
| Max | 1.00 | 0.813 | 1.00 |

| Parameter | Estimate | Estimated Error | Lower 95% Credible Interval | Upper 95% Credible Interval | Rhat | Bulk ESS | Tail ESS |
|---|---|---|---|---|---|---|---|
| Intercept | 1.92 | 0.23 | 1.48 | 2.38 | 1 | 11639 | 23637 |
| Arm | -1.90 | 0.29 | -2.49 | -1.37 | 1 | 13842 | 25773 |
| Baseline | 0.02 | 0.01 | 0 | 0.04 | 1 | 19943 | 35506 |
| Standard deviation of random intercept | 0.99 | 0.12 | 0.78 | 1.25 | 1 | 15625 | 30961 |

| Date of Data Transfer | Number of Participants Analysed (Delayed arm, Immediate arm) | Bayes Factors | |
|---|---|---|---|
| | | Testing for evidence against no benefit of intervention (evidence against no benefit) | Testing for positive treatment effect of the intervention (evidence in favour of positive effect) |
| 11/24/2021 | 20 (7,13) | 59.8 | 1.27 |
| 12/15/2021 | 23 (9,14) | 378 | 6.44 |
| 12/22/2021 | 29 (13, 16) | 2940 | 33.7 |
| 1/5/2022 | 37 (16,21) | 277 000 | 2340 |
| 1/12/2022 | 41 (17,24) | 2 500 000 | 19 500 |
| 1/26/2022 | 45 (20, 25) | 1 590 000 | 14 700 |
| 5/8/2022 | 75 (39, 36) | 177 000 000 | 1 250 000 |

| Serious Adverse Events Reported | Delayed arm (n) | Immediate arm (n) | Outcome |
|---|---|---|---|
| **Self-Reported Symptom** | | | |
| Admitted to hospital due to a ‘chest infection with reduced foetal movement’ | 0 | 1 | Participation in the study was paused |
| **Total Number of Serious Adverse Events** | **0** | **1** | |

| Adverse Events Reported | Delayed arm (n) | Immediate arm (n) |
|---|---|---|
| **Self-Reported Symptoms** | | |
| Spinal procedure | 1 | 0 |
| Recent leg surgery | 0 | 1 |
| Deranged bloods | 1 | 0 |
| Joint pain | 0 | 2 |
| Headache | 1 | 0 |
| Worsening of a gynae issue | 0 | 1 |
| Endometriosis | 0 | 1 |
| Breast lump | 1 | 0 |
| Menopause symptoms | 1 | 0 |
| Cardiac issues | 1 | 0 |
| Viral infection (NOS) | 1 | 1 |
| Acute gastro-enteritis | 0 | 1 |
| Chest infection and tonsilitis | 0 | 1 |
| Contracted COVID | 4 | 2 |
| Respiratory virus (NOS) | 1 | 0 |
| Chest infection | 1 | 0 |
| Worsened mental health | 1 | 0 |
| Suicidal ideation | 0 | 1 |
| **Responses to Changes to Health and Work questionnaire –** *Have you received any new treatments?* | | |
| Propanol | 1 | 1 |
| Fluoxetine | 1 | 0 |
| Anxiety medication (NOS) | 1 | 0 |
| Amitriptyline | 1 | 0 |
| Accessed counselling | 1 | 1 |
| **Total Number of Adverse Events** | **19** | **13** |

#### 7. FIGURE LEGENDS

**Supplementary Fig. 1. Primary Outcome Histograms**. Overlayed histograms which display the data by grouping data into "bins" of equal width along the x-axis. The histograms consist of a set of bars having bases on a horizontal axis (the x-axis) with centers at the bin midpoint and lengths equal to the bin interval sizes. In this case, the bin interval size was taken to be 1; hence the height of the bar (y-axis) centered at value *m* on the x-axis is equal to the count/frequency of participants who recorded *m* intrusive memories in the diary over the 7 day period.

**(A) Baseline measure for each arm.** Number of intrusive memories of traumatic events recorded by participants in a brief daily online intrusive memory diary for 7 days during the baseline week (i.e. run-in week) for both arms (red = delayed arm (control); n = 39: usual care for four weeks; blue = immediate arm; n = 36: immediate access to the intervention following the baseline week), showing that the two arms did not differ at baseline (i.e., before the intervention was provided to the immediate arm).

**(B) The primary outcome measure for each arm.** Number of intrusive memories of traumatic events recorded by participants in a brief daily online intrusive memory diary for 7 says during week 4 for both arms (red = delayed arm (control); n = 39: usual care for four weeks; blue = immediate arm; n = 36: immediate access to the intervention following the baseline week), showing that the immediate arm had fewer intrusive memories at week 4 compared to the delayed arm and that the number of intrusive memories for the immediate arm decreased between the baseline week and week 4

**Supplementary Fig. 2. Posterior Trace and Density Plots.** Posterior trace and density plots for MCMC Draws of fitted Bayesian model in final analyses. Figure made using *plot* function in the *brms* package. Referring to the final model parameters, the trace and density plots starting from the top and going down are for the population fixed effects of the intercept α, intevervention arm $\beta_1$, baseline number of intrusions $\beta_2$; and standard deviation of the random intercept.

**Supplementary Fig. 3. Posterior predictive checks.** Empirical cumulative distribution function for posterior predictive check of fitted Bayesian model in final analyses. The y-axis indicates the proportion of values falling below a value on the x-axis. The distribution function converges on 1.0 for values of 60, indicating that almost all observed counts were lower than that. The light blue lines labelled $y_{rep}$, represent the simulated data; the dark blue line labelled y, represents the actual data. We see that the dark blue line (the cumulative distribution function of the data) falls reasonably within the simulated data suggesting that the model could reasonably have generated the data.

**Supplementary Fig. 4. Posterior Density Plots for varying priors**. The figure combines a violin plot of the posterior density (colour coded by prior used), median, 66% and 95% quantile interval (in black) to give an "half eye plot" of the posterior. The y-axis on each plot represents the different priors used: student-t distribution (3 degrees of freedom, location 0, scale 2), Normal (mean 0, SD 2) , Normal (mean 0, SD 10), and Normal (mean 0, SD 20).

**(A)** Posterior Density Plots for the population level effects of intervention arm

**(B)** Posterior Density Plots for the population level effects of baseline number of intrusions.

**Supplementary Fig. 5. Posterior Density Plots for varying models.** The figure combines a violin plot of the posterior density (colour coded by model used), median, 66% and 95% quantile interval (in black) to give an "half eye plot" of the posterior. The y-axis on each plot represents the model used: Zero Inflated Poisson, Zero Inflated Negative Binomial, Poisson with Random Intercept, Poisson and Negative Binomial.

**(A)** Posterior Density Plots for the population level effects of intervention arm

**(B)** Posterior Density Plots for the population level effects of baseline number of intrusions.

**Supplementary Fig. 6. Posterior Density Plots for data with and without outliers.** The figure combines a violin plot of the posterior density (colour coded whether outliers were included or removed), median, 66% and 95% quantile interval (in black) to give an “half eye plot” of the posterior. The y-axis on each plot represents that the model was fitted using data without outliers and data with outliers.

**(A)** Posterior Density Plots for the population level effects of intervention arm

**(B)** Posterior Density Plots for the population level effects of baseline number of intrusions.

**Supplementary Fig. 7. Posterior Density Plots for all data and non-imputed data.** The figure combines a violin plot of the posterior density (colour coded whether all data was used, or only non-imputed data was used), median, 66% and 95% quantile interval (in black) to give an “half eye plot” of the posterior. The y-axis on each plot represents that the model was fitted using data without outliers and data with outliers.

**(A)** Posterior Density Plots for the population level effects of intervention arm

**(B)** Posterior Density Plots for the population level effects of baseline number of intrusions.

**Supplementary Fig. 8. Bayes Factor vs Sample Size plot to test for a positive treatment effect under the optimised intervention.** Bayes Factor (along y-axis) and sample size (along x-axis) graph made using the *super_bf* function in *bayesmedr* package. The mean of the primary outcome for the immediate intervention arm under optimsied intervention is varying (lines colour coded by mean), and data from interim analyses 4 was used for the mean and

SD of the delayed intervention arm, and the SD of the immediate intervention arm. The x-axis represents the total sample size (with roughly equal split between arms).

**Supplementary Fig. 9. Bayes Factor vs Sample Size plot to compare the optimised intervention to the former un-optimised intervention.** Bayes Factor (along y-axis) and sample size (along x-axis) graph made using the *infer_bf* and *equiv_bf* functions in *bayesmedr* package. The mean of the primary outcome for the immediate intervention arm under the optimised intervention is varying (lines colour coded by mean), and data from fourth interim Bayesian analysis was used for the mean and SD of the immediate intervention arm under un-optimised intervention, and the SD of the immediate intervention arm under optimised intervention. The x-axis represents the sample size of the immediate arm under the optimised intervention.

**(A)** Bayes Factor vs Sample Size plot to test for equivalence

**(B)** Bayes Factor vs Sample Size plot to test for non-inferiority

#### 8. FIGURES

Supplementary Fig. 1 through 9 are provided below in numerical order.

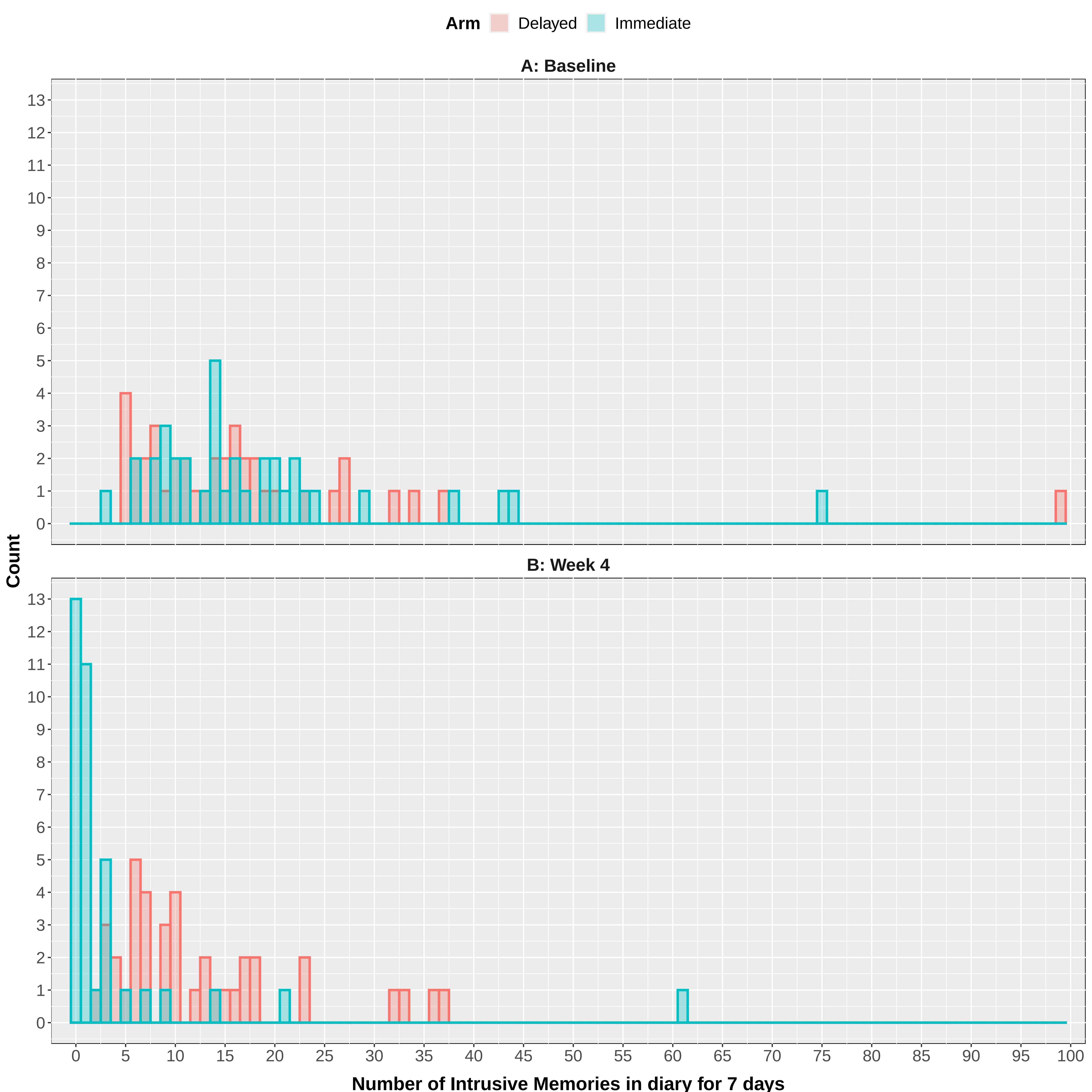

Arm
Delayed
Immediate
A: Baseline
B: Week 4
Count
Number of Intrusive Memories in diary for 7 days

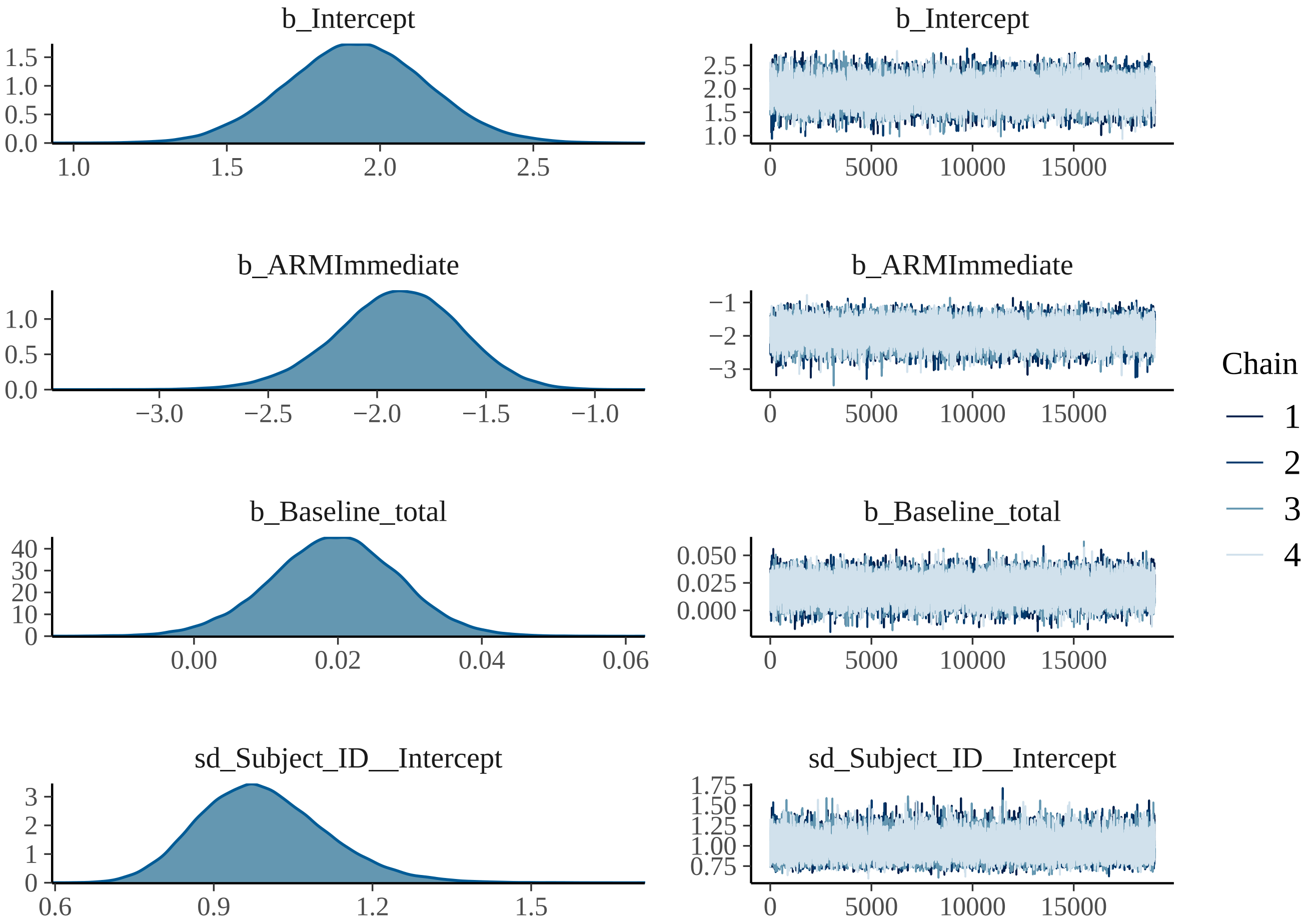

b_Intercept
b_ARMImmediate
b_Baseline_total
sd_Subject_ID__Intercept
Chain
1
2
3
4
0
5000
10000
15000

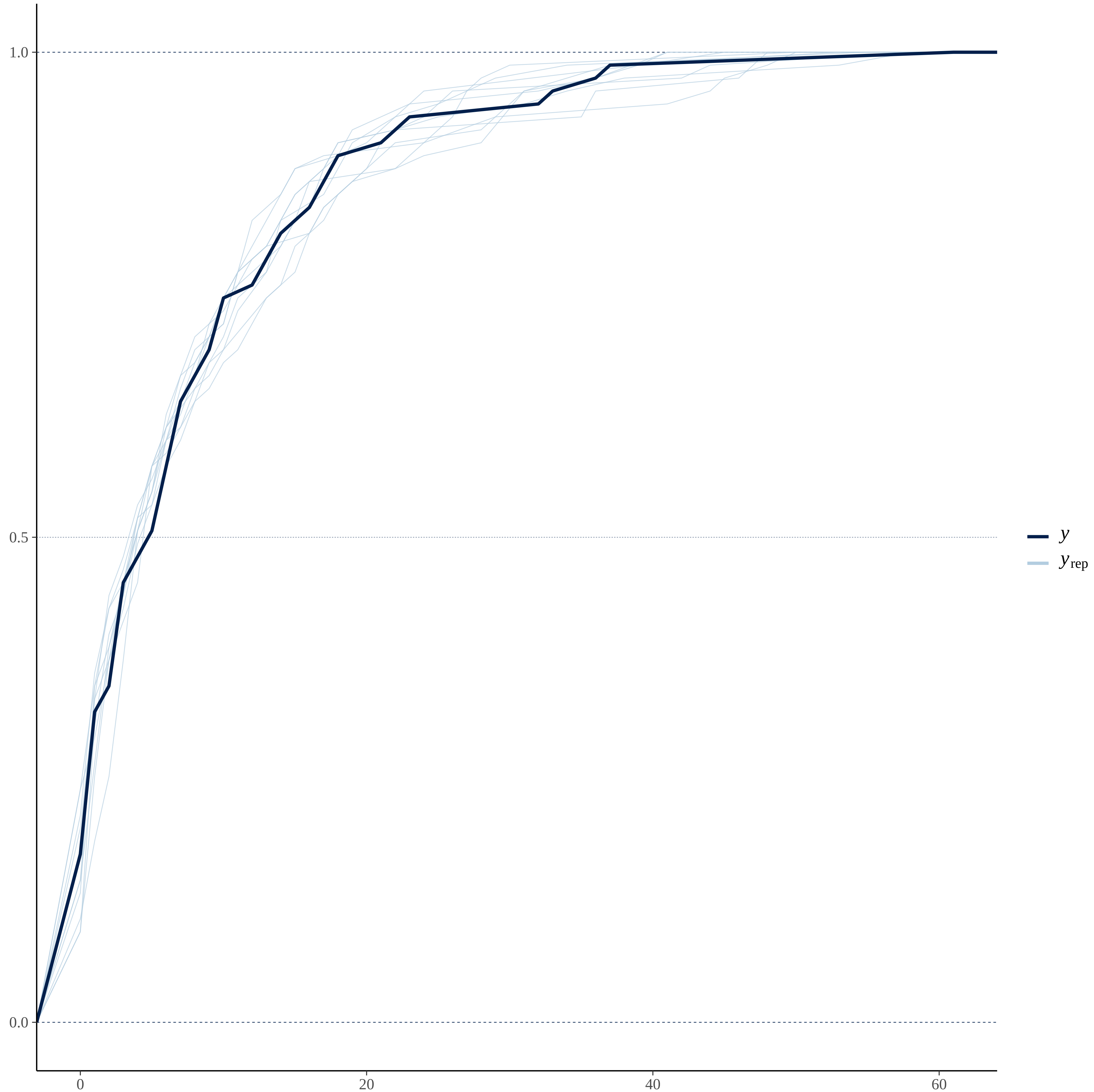

1.0
0.5
0.0
0
20
40
60
y
y rep

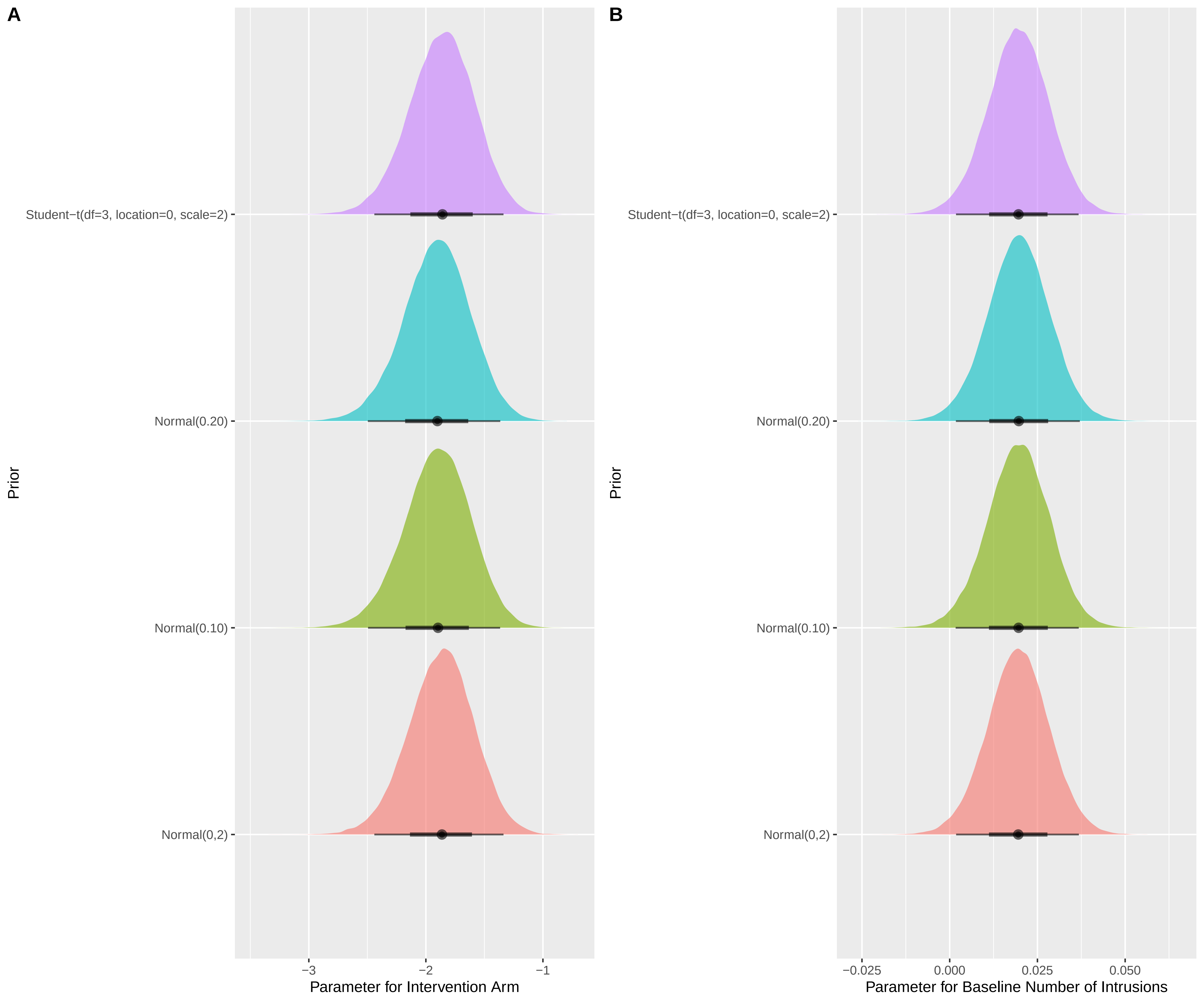

A
B
Student−t(df=3, location=0, scale=2)
Normal(0.20)
Normal(0.10)
Normal(0,2)
Prior
−3
−2
−1
Parameter for Intervention Arm
−0.025
0.000
0.025
0.050
Parameter for Baseline Number of Intrusions

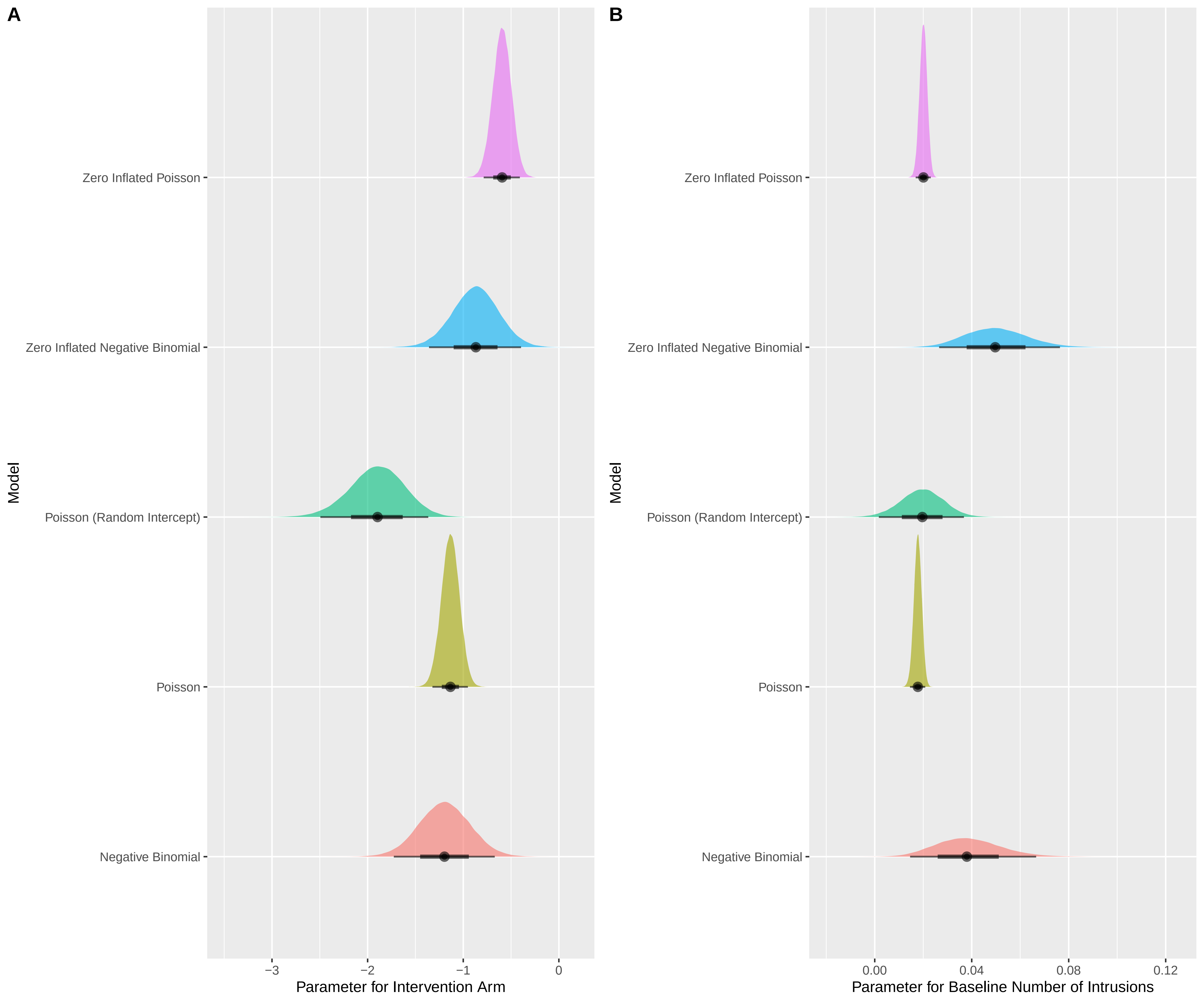

A
Zero Inflated Poisson
Zero Inflated Negative Binomial
Poisson (Random Intercept)
Poisson
Negative Binomial
Model
−3
−2
−1
0
Parameter for Intervention Arm
B
Zero Inflated Poisson
Zero Inflated Negative Binomial
Poisson (Random Intercept)
Poisson
Negative Binomial
Model
0.00
0.04
0.08
0.12
Parameter for Baseline Number of Intrusions

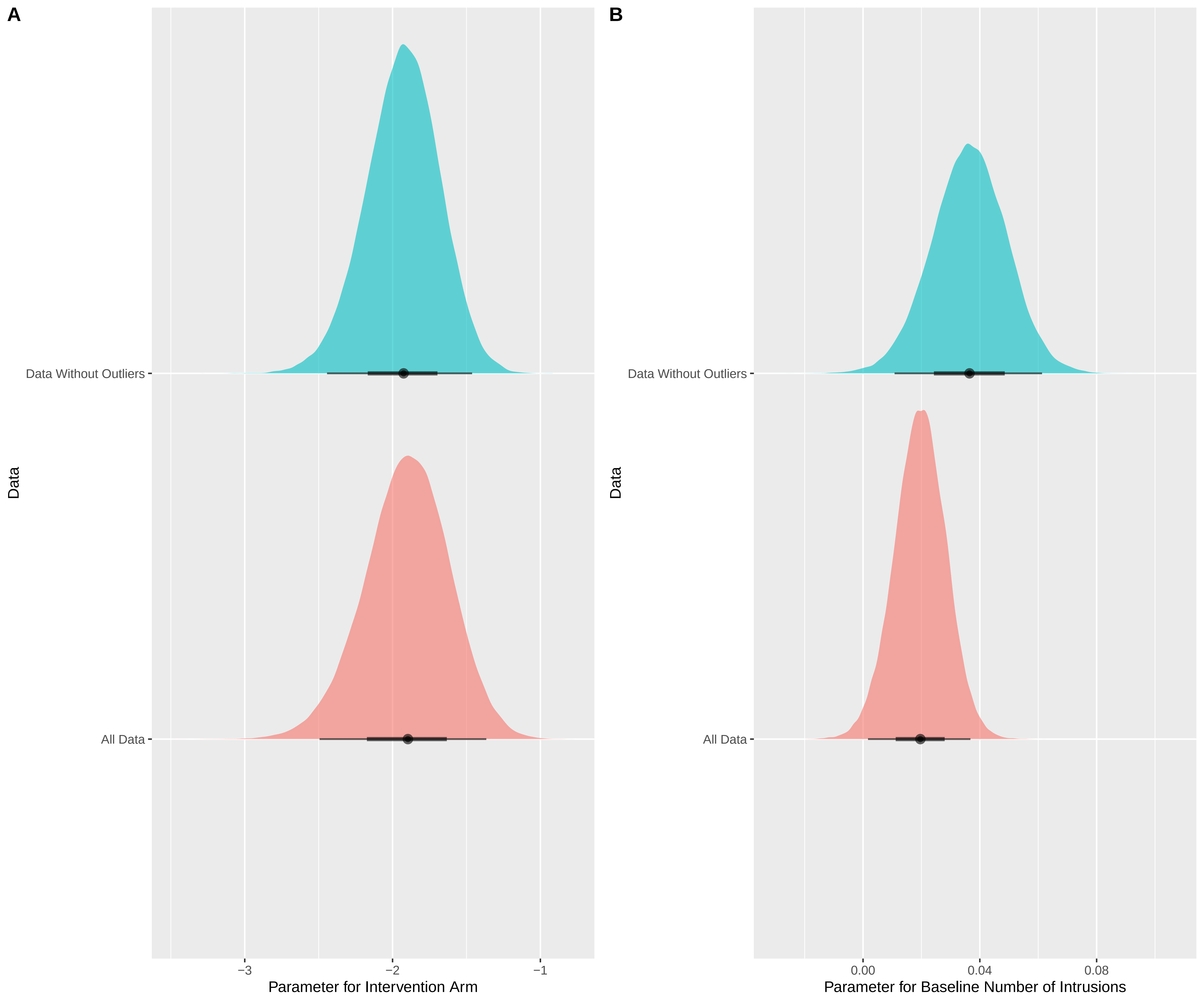
A
B
Data Without Outliers
All Data
Data
−3
−2
−1
Parameter for Intervention Arm
0.00
0.04
0.08
Parameter for Baseline Number of Intrusions

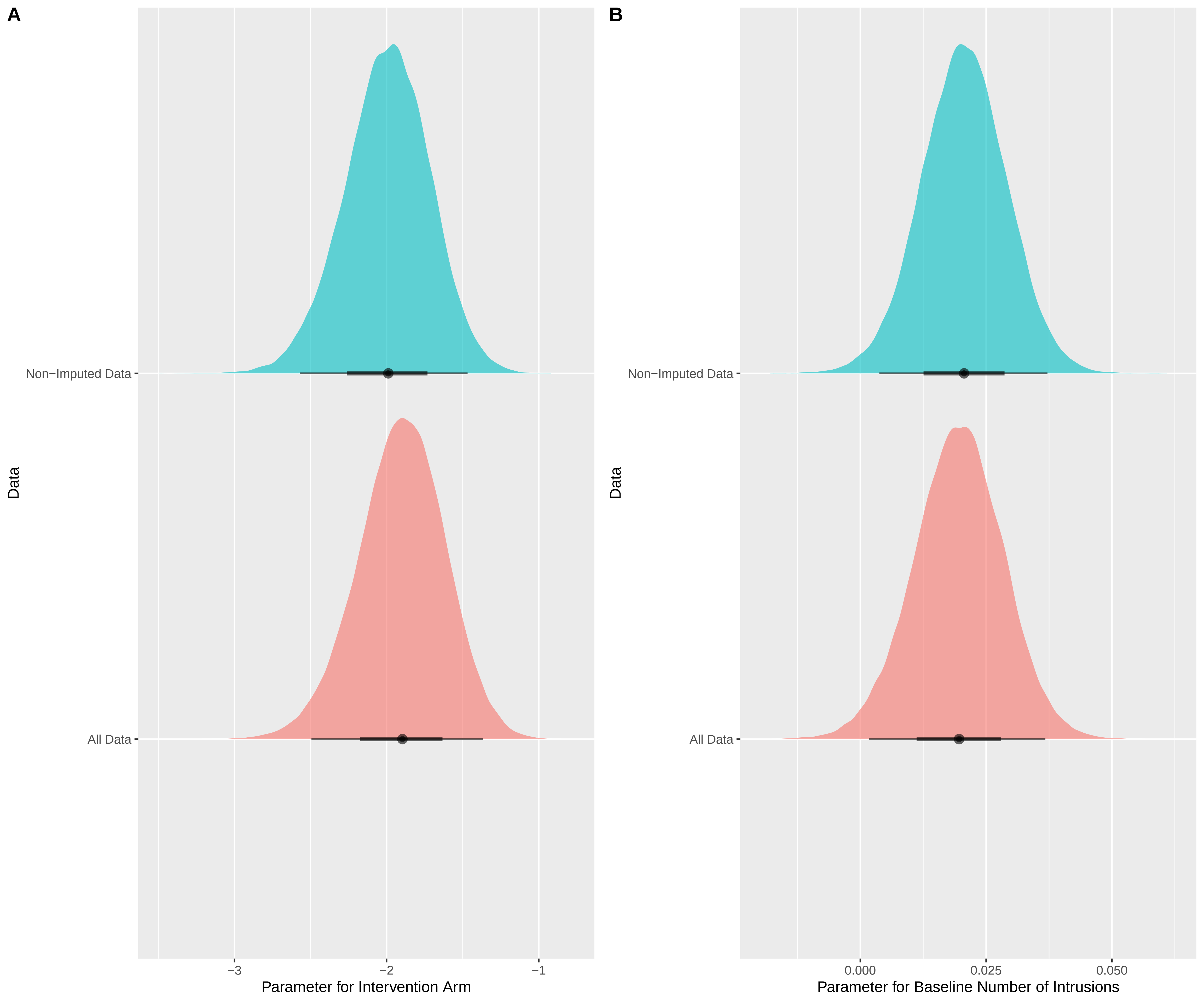
A
Non−Imputed Data
All Data
Data
−3
−2
−1
Parameter for Intervention Arm
B
Non−Imputed Data
All Data
Data
0.000
0.025
0.050
Parameter for Baseline Number of Intrusions

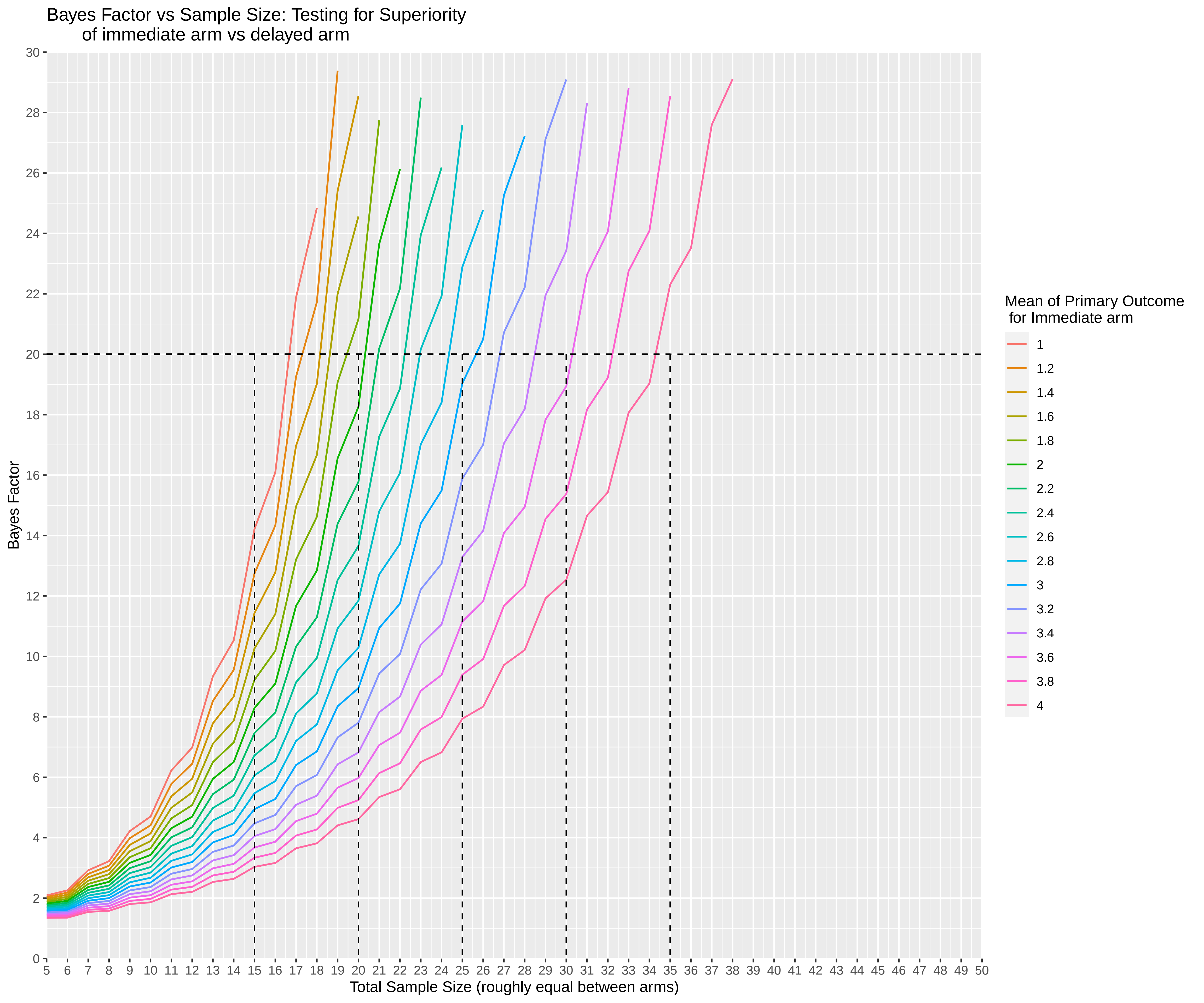
Bayes Factor vs Sample Size: Testing for Superiority
of immediate arm vs delayed arm
Bayes Factor
Total Sample Size (roughly equal between arms)
0
2
4
6
8
10
12
14
16
18
20
22
24
26
28
30
5 6 7 8 9 10 11 12 13 14 15 16 17 18 19 20 21 22 23 24 25 26 27 28 29 30 31 32 33 34 35 36 37 38 39 40 41 42 43 44 45 46 47 48 49 50
Mean of Primary Outcome
for Immediate arm
1
1.2
1.4
1.6
1.8
2
2.2
2.4
2.6
2.8
3
3.2
3.4
3.6
3.8
4

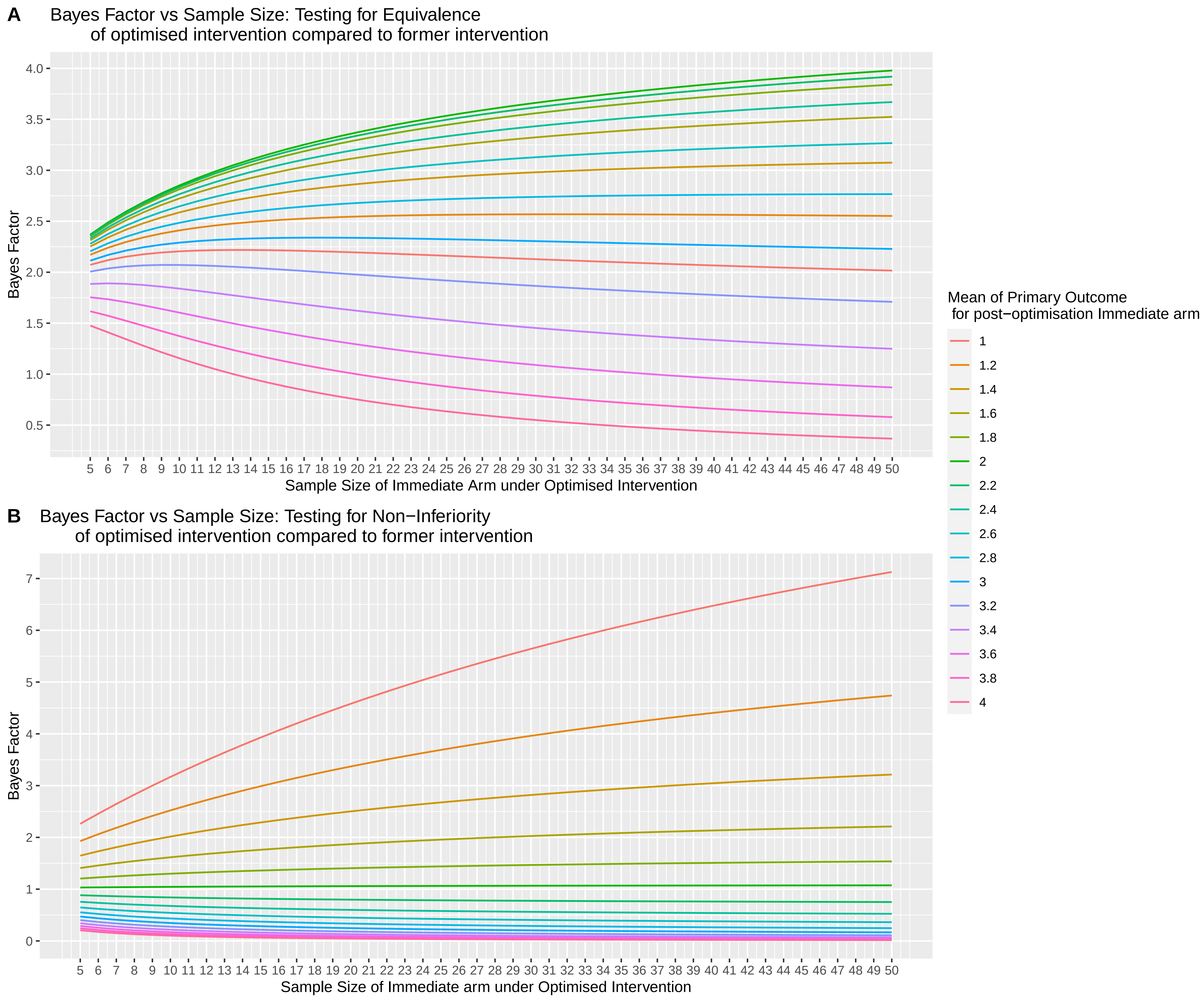
A
Bayes Factor vs Sample Size: Testing for Equivalence of optimised intervention compared to former intervention
Bayes Factor
Sample Size of Immediate Arm under Optimised Intervention
B
Bayes Factor vs Sample Size: Testing for Non−Inferiority of optimised intervention compared to former intervention
Bayes Factor
Sample Size of Immediate arm under Optimised Intervention
Mean of Primary Outcome for post−optimisation Immediate arm
1
1.2
1.4
1.6
1.8
2
2.2
2.4
2.6
2.8
3
3.2
3.4
3.6
3.8
4
